# Postnatal Brain Magnetic Resonance Imaging Trajectories and Maternal Intelligence Predict Neurodevelopmental Outcomes in Complex Congenital Heart Disease

**DOI:** 10.1101/2023.10.11.23296856

**Authors:** Vincent K. Lee, Rafael Ceschin, William T. Reynolds, Benjamin Meyers, Julia Wallace, Douglas Landsittel, Heather M. Joseph, Daryaneh Badaly, J. William Gaynor, Daniel Licht, Nathaniel H. Greene, Ken M. Brady, Jill V. Hunter, Zili D. Chu, Elisabeth A. Wilde, R. Blaine Easley, Dean Andropoulos, Ashok Panigrahy

**Affiliations:** Department of Bioengineering, University of Pittsburgh, Pittsburgh, Pennsylvania, USA; Department of Radiology, University of Pittsburgh, Pittsburgh, Pennsylvania, USA; Department of Biomedical Informatics, University of Pittsburgh, Pittsburgh, Pennsylvania, USA; Department of Epidemiology and Biostatistics, Indiana University School of Public Health-Bloomington, Bloomington, Indiana, USA; Department of Psychiatry, University of Pittsburgh, Pittsburgh, PA; Learning and Development Center, Child Mind Institute, New York, New York, USA; Division of Cardiothoracic Surgery, Children’s Hospital of Philadelphia, Philadelphia, Pennsylvania, USA; Division of Neurology, Children’s Hospital of Philadelphia, Philadelphia, Pennsylvania, USA; Department of Pediatric Anesthesiology, Baylor College of Medicine, Houston, TX Anesthesiology, Oregon Health and Sciences University, Portland, Oregon, USA Anesthesiology, Lurie Children’s Hospital, Northwestern University, Chicago, Illinois, USA; Department of Radiology, Baylor College of Medicine, Houston, TX; H. Ben Taub Department of Physical Medicine and Rehabilitation, Baylor College of Medicine, Houston, Texas, USA, and TX, Department of Neurology, University of Utah School of Medicine, Salt Lake City, UT, USA; Department of Anesthesiology, Perioperative and Pain Medicine, Texas Children’s Hospital, Houston, Texas, USA

**Author notes:** Address Correspondence and Reprint Request to: Ashok Panigrahy, MD, Department of Pediatric Radiology, Children’s Hospital of Pittsburgh of UPMC, 4401 Penn Avenue, Pittsburgh, PA 15224. these primary co-authors contributed equally (Vincent K. Lee, Rafael Ceschin). these senior co-authors contributed equally (Dean Andropoulos, Ashok Panigrahy).

## Abstract

**Importance:** Congenital heart disease (CHD), especially the complex forms – such as hypoplastic left heart syndrome (HLHS) and transposition of the great arteries (TGA) – have been linked to neurodevelopmental deficits including impairments in gross cognitive functions, language abilities, and visuo-motor skills. The prognostic value of early infant brain trajectories and cumulative impact of demographic factors in relation to childhood neurodevelopmental outcomes is unknown.

**Objective:** To determine whether early structural brain trajectories predict early childhood neurodevelopmental deficits in complex CHD patients and to assess relative cumulative risk profiles of clinical, genetic, and demographic risk factors across early development.

**Design, Setting, and Participants:** We studied a prospective cohort study of term neonates with complex CHD (TGA and HLHS) were recruited at Texas Children’s Hospital between 2005-2011. Participants underwent structural MRI scans at three time points (one preoperative scan, one postoperative scan within 7 days of surgery, and one follow-up postoperative scan at 4 months). Participants also received three neurodevelopmental assessments at 1, 3, and 5 years of age.

**Main Outcomes and Measures:** Brain region volumes (macrostructure) and white matter tract (microstructure) fractional anisotropy (FA) and radial diffusivity (RD) were measured from the MRI scans acquired in the three neonatal time points. Three imaging trajectories – changes in volume, FA and RD, over time – corresponding to periods of brain changes were determined: perioperative (preoperative to postoperative #1), post-surgical (postoperative #1 to postoperative #2), and overall (preoperative #1 to postoperative #2). Gross cognitive, language, and visuo-motor outcomes were assessed with the Bayley Scales of Infant and Toddler Development, Third Edition (Bayley-III) at 1 and 3 years, and with the Wechsler Preschool and Primary Scale of Intelligence, Third Edition (WPPSI-III Full-Scale IQ and Verbal IQ, and Beery-Buktenica Developmental Test of Visual-Motor Integration (VMI)., 6^th^ Edition at 5 years. The analysis included development of predictive multi-variable models incorporating other known risk factors (i.e., heart lesion type, microdeletion-related genetic abnormality, and maternal IQ) of poor neurodevelopmental outcomes in CHD.

**Results:** A total of 95 term (38.5±1.3 weeks gestational age) neonates with complex CHD (49 [51.6%] HLHS, 46 [48.4%] TGA; 42 [44.2%] girls) were analyzed. Reduced overall period trajectories predicted poor language outcomes: brainstem (p=0.0022) and white matter (p=0.0397) predicted poor 5-year verbal IQ; brainstem (p=0.0134), deep grey (p=0.0258), and FA of superior longitudinal fasciculus (SLF) (p=0.0256) predicted poor 3-year language; whole brain volume predicted poor performance on measures of language at 1 year. Maternal IQ was the strongest contributor to language outcome variance that increased from 37% at 1-year, up to 62% at 3-year, and up to 81% at 5-year testing. Genetic abnormality contribution to variance in these same models decreased from 41% in 1-year to about 25% at 3-year, and then to not significant in the 5-year assessments. Heart lesion type was found to be not significant in predicting outcomes in these models.

**Conclusion and Relevance:** A dysmaturation pattern of reduced postnatal trajectories of subcortical-cerebral white matter MRI metrics predicted poor early childhood neurodevelopmental outcomes, despite the high relative contribution of maternal IQ. Maternal IQ was cumulative over time, exceeding the influence of known innate cardiac and genetic factors in complex CHD, underscoring the importance of both heritable factors and parent-based environmental factors.

**Key Points:** *Question:* Do early infant brain trajectories in congenital heart disease (CHD) patients predict early childhood neurodevelopmental (ND) outcomes adjusted for known genetic abnormalities and maternal intelligence (IQ)?

*Findings:* Among infants with, reduced brainstem and white matter volumetric trajectories in children with CHD predicted language outcomes at five years, adjusting for maternal IQ and known genetic abnormalities. At the same time, known genetic abnormalities exerted a maximum effect at 1-year relative to 5-year neurodevelopmental testing. Maternal IQ was the most substantial contributor to ND outcome variance, nearly doubling from 1-year relative to 5-year time points.

*Meaning:* Postnatal infant brain trajectories may aid in the prognostication of early childhood neurodevelopment outcomes in complex CHD. The influence of maternal IQ is ***cumulative*** and can exceed the influence of known innate cardiac and genetic factors in complex CHD, underscoring the importance of not only heritable factors but also parent-based environmental factors.

## Introduction

Congenital heart disease (CHD) poses lifelong risks for complications, including neurodevelopmental deficits .^1,2^ Children with CHD experience heterogenous degrees of neurodevelopmental outcomes and trajectories, with language impairment the most prominent during early childhood.^3,4^ Given the persistent risk of neurodevelopmental deficits in individuals with CHD, it is imperative to evaluate long-term developmental trajectories (changes over time) to understand and potentially intervene in relation to their overall quality of life.

In individuals with CHD, alterations to the trajectory of brain development begin in utero, evolve over time, and are compounded by additional injury. These interact with a multitude of medical and demographic factors and lead to a heterogeneous array of adverse neurodevelopmental outcomes. Improved understanding of brain trajectories in the context of these factors can provides us with a conceptual framework for the development of targeted interventions that might improve outcomes. Cross-sectional studies of CHD patients in infancy have shown that delayed brain maturity^5–8^, reduced volumes^9–11^, and abnormality in connectivity^12–14^ are clearly associated with adverse neurodevelopment in CHD. Specifically, reduced total brain, brainstem, and white matter volumes have been shown to predict poor language outcomes at one year of age^15^. There are also a few studies examining brain growth trajectories between late fetal and neonatal periods^16–18^ pre-operative and post-operative time points^19^, and neonatal to infant periods showing some indication of reduced brain volume trajectory^11^. However, there is a paucity of trajectory studies assessing the predictive abilities of neonatal brain imaging in CHD for short- and long-term neurodevelopmental outcomes.

Past research, which has included extensive studies of multiple sources of exposures, has failed to adequately predict developmental delays, with significant unexplained (>60%)^2,4^ variance in developmental outcomes. It is well established that social-environmental factors such as parenting, maternal education and socio-economic status are strong contributors to infant and child development, and there is emerging evidence to suggest the effect is even greater in those infants and children with CHD. ^20–23^ Maternal intellectual ability, which has been poorly understood in CHD cohorts, also has direct influence on children’s intellectual development because it is a genetically based and heritable trait.^24^ Intellectual ability is also associated with adult life-course outcomes ^25^ that, in turn, shape the child-rearing environment.^26^

In this study, we primarily tested the hypotheses that reduced trajectory of early infant brain structures, as measured with multi-modal brain MRI, would be predictive of early childhood neurocognitive outcomes at five years of age, adjusting for maternal IQ, known genetic abnormalities (microdeletions), and other medical factors that are known to contribute to neurodevelopmental outcomes variance. We secondarily tested the hypotheses that postnatal trajectories of multi-modal brain structure and concomitant influence of demographic factors on early neurodevelopmental outcomes is *cumulative* (i.e., the strength of effect increases with increasing age) and could potentially exceed the influence of known innate clinical and genetic factors in CHD. As such, we also correlated the same early infant brain trajectories with comparable late infant (1-year) and toddler (3-year) neurodevelopmental outcomes within the same cohort, adjusting for similar covariates. Lastly, we also examined the predictive value of more perioperative and post-surgical brain trajectories over more specific time points in relation to comparable sequential neurodevelopmental domains adjusting for similar covariates in the same cohort.

## Methods

The study protocol, including all clinical data collection, acquisition of MRI data under sedation, and neurodevelopmental outcomes testing of subjects and maternal IQ testing was approved by the Baylor College of Medicine Institutional Review Board, and written informed parental consent was obtained. Written informed consent was obtained from a parent under Baylor College of Medicine IRB Protocols H-18531 and H-17115.

### Study Population

Neonates diagnosed with hypoplastic left heart syndrome (HLHS – requiring Norwood operation and Blalock-Taussig shunt placement), transposition of the great arteries (TGA) – requiring arterial switch operation or ventricular septal defect repair with aortic arch reconstruction were recruited into this study within the first 30 days of life at Texas Children’s Hospital from November 2005 to December 2011. Exclusion criteria were: 1) gestational age less than 35 weeks, 2) weight less than 2.0 kg at birth, 3) recognizable dysmorphic syndrome, and 4) preoperative cardiac arrest greater than 3 minutes.

### Magnetic Resonance Imaging Studies

The study design included three MRIs performed under anesthesia: 1) the preoperative scan acquired just prior to surgery, after tracheal intubation (39.42±1.53 weeks post-conceptual age at scan); 2) the post-operative #1 scan between 7-10 days post-operatively (40.66±1.53 weeks) under pentobarbital sedation; and 3) post-operative #2 scan between 4-6 months of life (61.34±7.51 weeks) under pentobarbital or propofol infusions. All MRI scans were acquired on the same 1.5T Intera scanner (Philips Medical Systems, Best, Netherlands). Each of the scans included volumetric 3-dimensional T1-weighted and diffusion imaging (15 gradient directions B=860s/mm). Detailed MRI sequence parameters are presented in the eMethods (eTable 1). All T1-weighted and diffusion images underwent in-house semi-automated segmentation or tractography pipelines as previously reported^6,27^ with summary descriptions provided in the eMethods. The following brain regions were included: cerebral cortex, cerebral white matter volume (WMV), brainstem, cerebellum, intracranial cerebrospinal fluid (CSF), deep grey matter (DGM), and whole brain with and without CSF. The following white matter tracts were included: genu, body, and splenium of the corpus callosum (CC); left and right cortico-spinal tract (CST-L and CST-R); left and right fronto-occipital fasciculus (FOF-L and FOF-R); left and right inferior longitudinal fasciculus (ILF-L and ILF-R); and left and right superior longitudinal fasciculus (SLF-L and SLF-R). The average fractional anisotropy (FA) and radial diffusivity (RD) values for each white matter tract were calculated.

### Trajectory of Structural Imaging Metrics

The trajectory of change in volume, FA, and RD were defined as change in imaging measurement (FA-index and mm^2^/s, respectively) over change in time between the two scans that encompass the trajectory period. The primary trajectory exposure for this study was the early infant trajectory period which was calculated spanning between preoperative to postoperative #2 scans. The secondary exposures were post-surgical trajectory (period between postoperative #1 and postoperative #2 scans) and perioperative trajectory (between preoperative and postoperative#1 scans). Because of the known dramatic differential increase in normative metrics in regional brain volume and DTI metrics during this period, no attempt was made to model all three time points into one trajectory measure.

### Neurodevelopmental Tests

The participants underwent neurodevelopmental assessment at one, three, and five years of age. At one and three years, the Bayley Scales of Infant and Toddler Development, 3^rd^ Edition (Bayley-III) was administered for indices of global cognitive, language, and motor functions. At 5 years, children completed the Wechsler Preschool and Primary Scale of Intelligence, 3^rd^ edition (WPPSI-III) and Beery-Buktenica Developmental Test of Visual-Motor Integration, 6^th^ edition (Beery VMI). To facilitate a conceptual comparison with 1- and 3-year outcomes, we focused our analyses on the WPPSI-III Full Scale IQ (as an index of global cognition), the WPPSI-III Verbal IQ (as an index of language-based skills), and the Beery VMI (as a measure of visual and motor skills). For each of the neuropsychological tests, norm-referenced scores were computed, comparing children to same-age peers.

### Non-imaging Factors

The non-imaging factors considered were factors previously recognized to be associated with neurodevelopmental outcomes in CHD: presence or absence of genetic abnormalities in general, and 22q deletion specifically; heart lesion type as single (HLHS) or double (TGA) ventricle; surgical variables – length of hospital stay, whether the participant had open heart surgery; demographic factors – SES status (assessed with Hollingshead Four-Factor Index of Socioeconomic Status); maternal IQ (assessed with Wechsler Abbreviated Scale of Intelligence, Second Edition; WASI-II); sex; race; ethnicity; birth weight; and white matter injury (WMI).

### Statistical Analysis

Statistical analysis was performed using SAS software, version 9.4 (SAS Institute Inc.). A priori, we determined that the most relevant comparison was between the early infant imaging trajectory, which spans the longest duration between scans, and 5-year outcome, which represents the farthest neurodevelopmental assessment in this study. To assess whether brain imaging trajectories – and other factors that might impact neurodevelopment – predict neurodevelopmental outcomes, a multi-variable model was developed. This model examined the contribution of each imaging trajectory to each neurodevelopmental outcome while incorporating a set of non-imaging risk factors likely to contribute to the neuropsychological assessment variability as covariates. The covariate selection involves further refinement of risk factors demonstrating significant associations (alpha <0.05) with neurodevelopmental tests, from an initial regression analysis, through collinearity tests and screening variable elimination. Consequently, maternal IQ, genetic abnormality, and cardiac ventricles to model HLHS-TGA status were incorporated into the final model. The covariate selection process is further detailed in the eMethods. Post-hoc factor contribution analysis was conducted for each multi-variable regression test that demonstrated significant association between imaging trajectory and neuropsychological test. This analysis examined the contributions of each independent variable – imaging as well as non-imaging in the final multi-variable model – to the variance in test performance.

## Results

The study cohort was 95 (42 [44.2%] female) neonatal patients (mean gestational age at birth of 38.23±1.23 weeks) with demographics provided in Table 1. Due to attrition in imaging and follow-up neurodevelopmental assessments, the participant numbers in our multi-variable models were reduced. Our primary multi-variable models comparing early infant trajectory imaging to neurodevelopmental outcomes had: 54 (23 [42.6%] female) patients at 1-year, 39 (15 [38.5%] female) at 3-year, and 36 (16 [44.4%] female) at 5-year assessments. Our secondary models comparing post-surgical trajectory imaging to neurodevelopmental outcomes had 50 (22 [44.0%] female) patients at 1-year, 37 (16 [43.2%] female) at 3-year, and 36 (16 [44.4%] female) at 5-year assessments. The secondary models comparing perioperative trajectory imaging to neurodevelopmental outcomes had 48 (20 [41.7%] female) patients at 1-year, 36 (13 [36.1%] female) at 3-year, and 36 (15 [41.7%] female) at 5-year assessments. Detailed cohort information and summary statistics are provided in eTable 3a-c. WMI incidence and volumes were not predictive of neurodevelopmental outcomes at any time points (see eResults).

**Table 1.** Demographics and Risk Factors.

|  | Recruited Cohort<br>(N=95) | 1-Year Cohort<br>(N=54) | 3-Year Cohort<br>(N=39) | 5-Year Cohort<br>(N=36) |
| --- | --- | --- | --- | --- |
| Parental SES | 40.66±14.91 | 39.78±14.52 | 41.89±13.78 | 41.30±13.63 |
| Maternal IQ | 102.92±16.73 | 102.5±17.01 | 104.90±16.87 | 104.11±18.17 |
| Birth Weight (g) | 3157±489 | 3186±513 | 3164±462 | 3190±487 |
| Gestational Age at Birth<br>(Weeks) | 38.23±1.23 | 38.04±1.20 | 38.03±1.25 | 38.19±1.35 |
|  | Median (IQR) | Median (IQR) | Median (IQR) | Median (IQR) |
| Hospital Length of Stay<br>(days) | 26 (20-39) | 24 (20-37) | 23 (20-38) | 23 (20-31) |
| Open Sternum Duration<br>(days) | 2 (1-2) | 2 (1-2) | 2 (1-2) | 2 (1-2) |
|  | Incident (%) | Incident (%) | Incident (%) | Incident (%) |
| Female Sex | 42 (44.21%) | 23 (0.43%) | 15 (0.38%) | 16 (0.44%) |
| Race and Ethnicity |  |  |  |  |
| Ethnicity (Hispanic) | 33 (34.74) | 16 (0.3%) | 10 (0.26%) | 10 (0.28%) |
| Race (White) | 83 (87.37) | 48 (0.89%) | 37 (0.95%) | 34 (0.94%) |
| Race (Black) | 10 (10.53) | 6 (0.11%) | 2 (0.05%) | 2 (0.06%) |
| Race (Asian) | 2 (2.11) | 0 (0%) | 0 (0%) | 0 (0%) |
| Received Open<br>Sternum Surgery | 23 (24.21) | 10 (0.19%) | 10 (0.26%) | 8 (0.22%) |
| Any Genetic<br>Abnormality* | 19 (20) | 11 (0.2%) | 10 (0.26%) | 7 (0.19%) |
| 22q Deletion | 8 of 19 (8.42) | 5 of 11 (0.45%) | 5 of 10 (0.5%) | 4 of 7 (0.57%) |
| Cardiac Single<br>Ventricles | 49 (51.58) | 25 (0.46%) | 19 (0.49%) | 16 (0.44%) |
| White Matter Injury | 41 (43.62) | 34 (0.63%) | 27 (0.69%) | 23 (0.64%) |

\*Other genetic abnormalities: 4q-deletion, 19q-deletion, 14q13.1gain, xp22.31gain, 8p23.1gain, 7p21.3, 18q22.1gain, 12q24.2 duplication, 45XO, 9pq21.3gain, 9q.region-duplication

### Early Infant Brain Trajectories (Overall Period) and Childhood Outcomes

Reduced trajectory of brainstem (β=0.0976, confidence interval (CI) 95%, 0.0382-0.1571; p=0.0022; factor contribution (FC) of 32.1%) and WM volume (β=0.0023, CI 95%, 0.0001-0.0046; p=0.0397; FC 15.6%) predicted poor 5-year language performance (Table 2, Figure 2) in the final multi-variable model. Lower maternal IQ (β=0.5501, CI 95%, 0.3203-0.7798; p<0.0001; FC up to 80.7%) was also predictive of poor 5-year language performance. Reduced trajectory of brainstem (β=0.0711, CI 95%, 0.01574-0.1265; p=0.0134; FC 16.3%) and DGM (β=0.0085, CI 95%, 0.0011-0.0160; p=0.0258; FC 11.8%) volume predicted poor 3-year language performance in the final multi-variable model. In these analyses, lower maternal IQ (β=0.4013, CI 95%, 0.2323-0.5703; p<0.0001; FC up to 62.1%) and presence of genetic abnormality (β=-11.2008, CI 95%, -17.9216–-4.4800; p=0.002) also predicted poor language outcome. Additionally, reduced FA trajectory (β=2263.4210, CI 95%, 327.7828–4199.0590; p=0.0256; FC 21%) and increased RD trajectory (β=-6968840, CI 95%, -1306.54–-87.2294; p=0.0284; FC 21.4%) of the SLF-R predicted poor 3-year language performance in the final multi-variable model. In these analyses, presence of genetic abnormality predicted poor language (β=-17.0154, CI 95%, -25.3444–-8.86865; p=0.0011; FC up to 64%) performance as well. Besides language, increased RD trajectory (β=-779.745 CI 95%, -1510.71–-48.7761; p=0.0375; FC up to 9.7%) of the ILF-L predicted poor 3-year motor performance, which was also associated with lower maternal IQ (β=0.2779, CI 95%, 0.0956-0.4602; p=0.0042; FC 19.9%), presence of genetic abnormalities (β=-12.0905, CI 95%, -187026–-5.4785; p=0.0009; FC 28.6%), and single ventricle status (β=12.9712, CI 95%, 7.1091-18.8330; p=0.0001; FC 41.88%).

**Figure 1.**
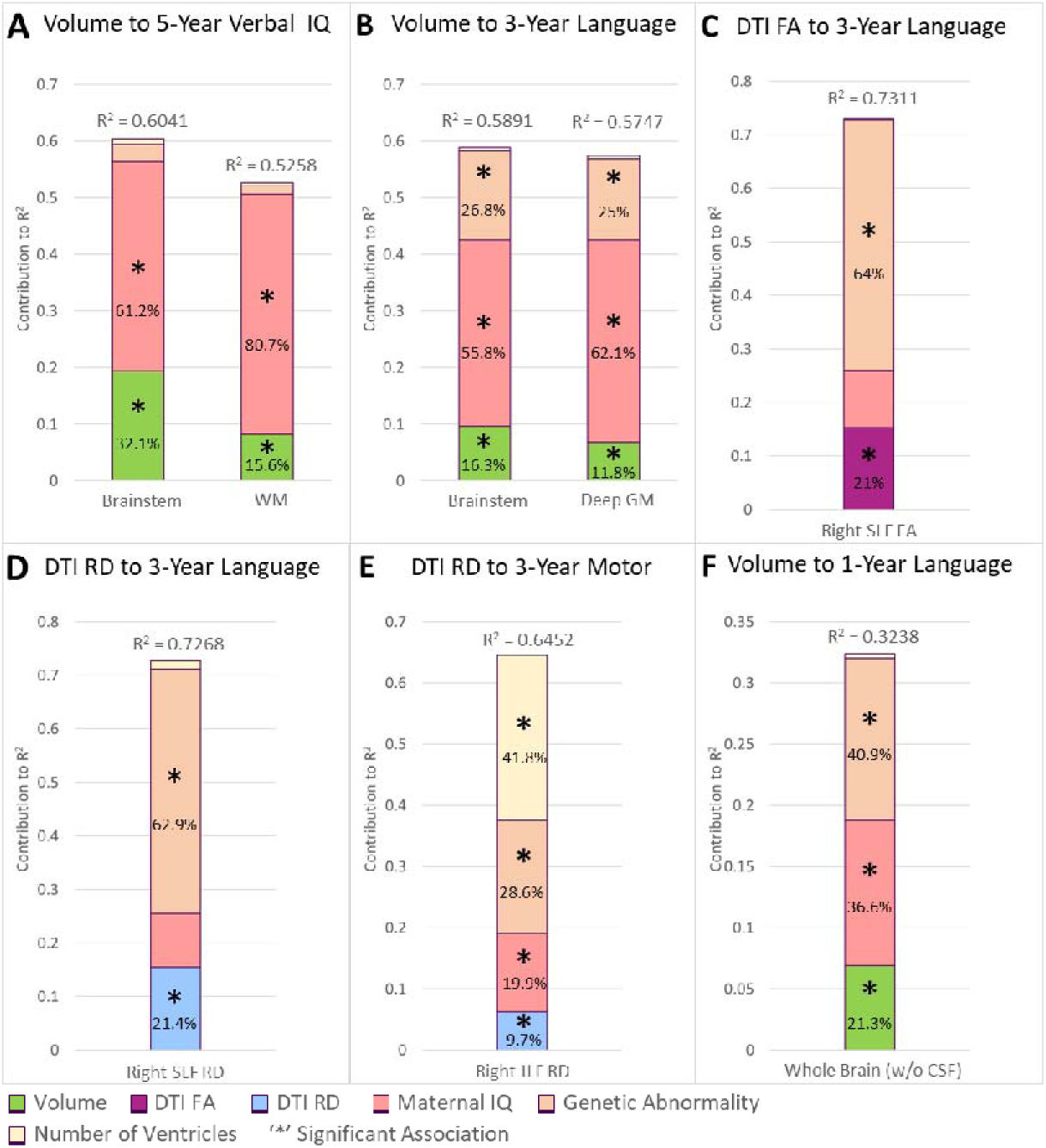
Early infant (Overall period) multi-modal imaging trajectories and non-imaging factors contribution to variability in early childhood (A-E) and infant (F) outcomes. Reduced early infant (overall period) volume trajectories predicted poor language outcome across all three neurodevelopmental outcome periods (5-year, 3-year, and 1-year outcomes). Altered WM microstructural early infant (overall period) trajectories in SLF predicted poorer performance on a measure of language and ILF predicted poor motor performance at the 3-year assessment. Macrostructure (volume) trajectories in this period predicted 1-year (up to 21% of variability) and early childhood (up to 32% of variability) performance variability in language/verbal outcomes. The only significant contribution of cardiac lesion type (number of ventricles) was to 3 year motor outcomes. As such, early infant neuroimaging trajectory variables showed remarkable consistency in the degree of ND outcome variance explained across 1-year, 3-year, 5-year time points. Genetic abnormality contribution to outcome relatively deceased from 3-year to 5-year time points. In contrast, maternal IQ contributed to the outcomes, and the contribution to variability clearly showed a marked cumulative increase over time from the 1-year assessment (37%) to early childhood (up to 81%).

**Figure 2.**
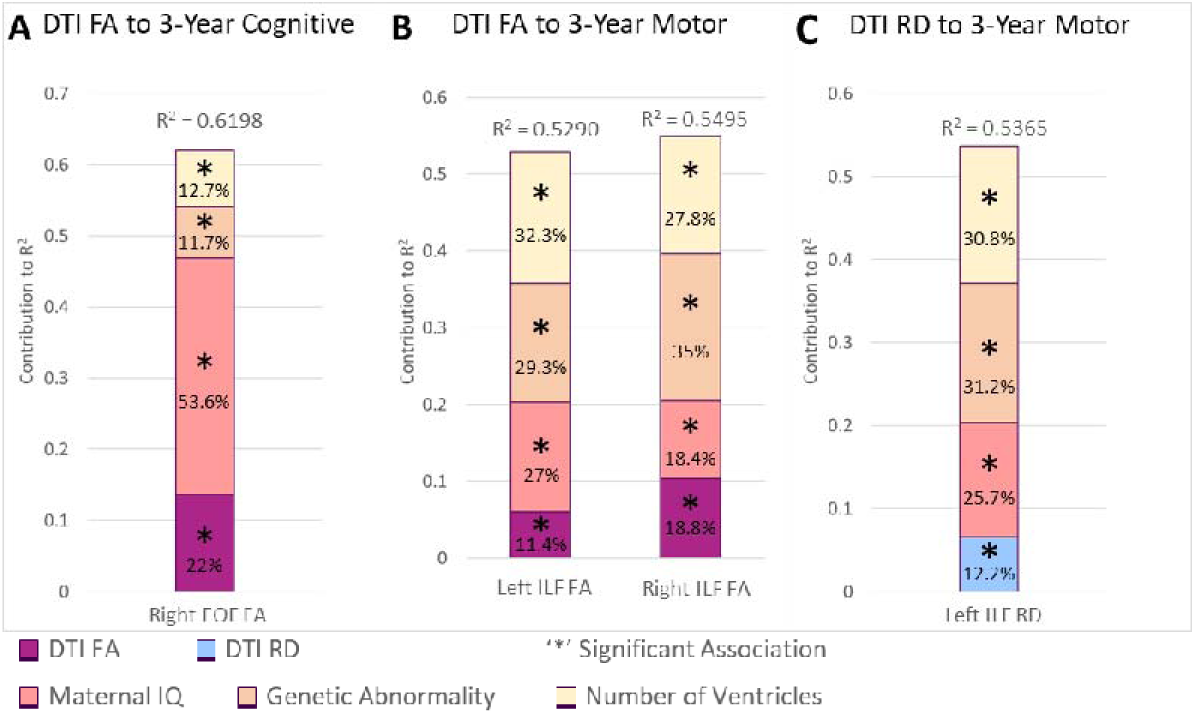
Post-surgical period imaging trajectories and non-imaging factors contribution to variability in early childhood outcomes. The post-surgical period imaging trajectories did not predict 5-year or 1-year outcomes. Volume trajectories from this period had no significant associations with any neurodevelopmental outcomes. Microstructure (FA and RD from diffusion imaging) accounted for up to 22% of cognitive and 19% of motor variability in 3-year outcomes. Maternal IQ contributed up to 66% in cognitive and 27% motor outcomes at 3 years. Genetic abnormality and number of cardiac ventricles were also co-contributors to 3 year outcome variability for this period.

**Table 2.** Adjusted Regression Analysis of Multi-Modal Imaging Trajectories to Neurodevelopmental Outcomes.

| Neurodevelopmental Outcomes | Imaging Biomarker | R^2 | Imaging Trajectory |  | Maternal IQ |  | Genetic Abnormality |  | Cardiac Ventricles |  |
| --- | --- | --- | --- | --- | --- | --- | --- | --- | --- | --- |
|  |  |  | β Coefficient (95% CI) | p-value | β Coefficient (95% CI) | p-value | β Coefficient (95% CI) | p-value | β Coefficient (95% CI) | p-value |
|  | Early Infant Volume Trajectory |  |  |  |  |  |  |  |  |  |
| 5 Year |  |  |  |  |  |  |  |  |  |  |
| WPPSI-III Verbal IQ | Brainstem | 0.6041 | 0.0977 (0.0382 - 0.1571) | 0.0022 | 0.483 (0.27 - 0.696) | <.0001 | -5.26 (-13.31 - 2.78) | 0.1915 | 2.42 (-4.16 - 8.99) | 0.4582 |
| WPPSI-III Verbal IQ | WMV | 0.5258 | 0.0023 (0.0001 - 0.0046) | 0.0397 | 0.55 (0.32 - 0.78) | <.0001 | -3.91 (-12.79 - 4.98) | 0.3762 | 2 (-5.39 - 9.38) | 0.5845 |
| 3 Year |  |  |  |  |  |  |  |  |  |  |
| Bayley-III Language | Brainstem | 0.5891 | 0.0711 (0.0157 - 0.1265) | 0.0134 | 0.401 (0.232 - 0.57) | <.0001 | -10.9 (-17.53 - -4.27) | 0.002 | 1.92 (-3.86 - 7.71) | 0.504 |
| Bayley-III Language | DGM | 0.5747 | 0.0085 (0.0011 - 0.016) | 0.0258 | 0.45 (0.279 - 0.622) | <.0001 | -11.2 (-17.92 - -4.48) | 0.0018 | 2.03 (-3.86 - 7.91) | 0.4888 |
| 1 Year |  |  |  |  |  |  |  |  |  |  |
| Bayley-III Language | Whole Brain | 0.3238 | 0.0008 (0.0001 - 0.0016) | 0.0286 | 0.257 (0.082 - 0.432) | 0.0048 | -11.49 (-18.89 - -4.1) | 0.003 | 1.6 (-4.48 - 7.68) | 0.5991 |
|  | Post-Surgical Volume Trajectory - No Significant Findings |  |  |  |  |  |  |  |  |  |
|  | Perioperative Volume Trajectory |  |  |  |  |  |  |  |  |  |
| 3 Year |  |  |  |  |  |  |  |  |  |  |
| Bayley-III Language | Brainstem | 0.7398 | 0.0073 (0.0019 - 0.0126) | 0.0111 | 0.419 (0.217 - 0.62) | 0.0004 | -6.05 (-11.92 - -0.18) | 0.044 | 0.52 (-4.3 - 5.35) | 0.8218 |
| Bayley-III Motor | CSF | 0.5918 | -0.0001 (-0.0003 - 0) | 0.0365 | 0.21 (-0.084 - 0.503) | 0.1495 | -13.59 (-22.35 - -4.84) | 0.0044 | 11.78 (4.68 - 18.88) | 0.0027 |
| 1 Year |  |  |  |  |  |  |  |  |  |  |
| Bayley-III Cognitive | CSF | 0.4949 | -0.0002 (-0.0003 - -0.0001) | 0.0062 | 0.2 (-0.056 - 0.457) | 0.1193 | -2.54 (-11.66 - 6.58) | 0.5698 | 5 (-1.93 - 11.92) | 0.1488 |
|  | Early Infant DTI FA Trajectory |  |  |  |  |  |  |  |  |  |
| 3 Year |  |  |  |  |  |  |  |  |  |  |
| Bayley-III Language | SLF-R | 0.7311 | 2263 (328 - 4199) | 0.0256 | 0.188 (-0.005 - 0.381) | 0.0556 | -17.02 (-25.34 - -8.69) | 0.0008 | 1.14 (-5.11 - 7.38) | 0.6985 |
|  | Post-Surgical DTI FA Trajectory |  |  |  |  |  |  |  |  |  |
| 3 Year |  |  |  |  |  |  |  |  |  |  |
| Bayley-III Cognitive | FOF-R | 0.6198 | 3769 (1055 - 6483) | 0.0082 | 0.351 (0.189 - 0.513) | 0.0001 | -6.44 (-12.79 - -0.09) | 0.0472 | 5.86 (0.3 - 11.43) | 0.0397 |
| Bayley-III Motor | ILF-L | 0.529 | 2760 (201 - 5319) | 0.0354 | 0.321 (0.127 - 0.514) | 0.002 | -13.75 (-21.71 - -5.78) | 0.0014 | 12.83 (5.75 - 19.92) | 0.0009 |
| Bayley-III Motor | ILF-R | 0.5495 | 3178 (651 - 5705) | 0.0156 | 0.255 (0.05 - 0.46) | 0.0165 | -13.46 (-21.31 - -5.62) | 0.0015 | 10.65 (3.7 - 17.6) | 0.004 |
|  | Perioperative DTI FA Trajectory |  |  |  |  |  |  |  |  |  |
| 3 Year |  |  |  |  |  |  |  |  |  |  |
| Bayley-III Language | SLF-L | 0.8109 | 155 (46 - 263) | 0.009 | 0.503 (0.324 - 0.683) | <.0001 | -0.12 (-7.27 - 7.04) | 0.9721 | 2.95 (-1.41 - 7.31) | 0.166 |
| 1 Year |  |  |  |  |  |  |  |  |  |  |
| Bayley-III Cognitive | FOF-L | 0.3445 | 232 (11 - 452) | 0.0402 | 0.396 (0.168 - 0.624) | 0.0011 | -8.5 (-18.06 - 1.05) | 0.0796 | 2.19 (-5.68 - 10.07) | 0.576 |
| Bayley-III Cognitive | FOF-R | 0.401 | 331 (88 - 573) | 0.0089 | 0.375 (0.151 - 0.599) | 0.0017 | -9.07 (-19.07 - 0.92) | 0.0738 | 0.95 (-6.91 - 8.81) | 0.808 |
| Bayley-III Cognitive | ILF-L | 0.4085 | 326 (109 - 544) | 0.0043 | 0.384 (0.171 - 0.597) | 0.0008 | -7.73 (-16.82 - 1.36) | 0.0933 | 2.73 (-4.56 - 10.02) | 0.4537 |
| Bayley-III Cognitive | ILF-R | 0.3838 | 337 (81 - 593) | 0.0112 | 0.424 (0.191 - 0.658) | 0.0007 | -10.92 (-20.19 - -1.65) | 0.0222 | 2.99 (-4.69 - 10.67) | 0.4349 |
| Bayley-III Cognitive | SLF-L | 0.6266 | 257 (91 - 423) | 0.0042 | 0.665 (0.374 - 0.957) | 0.0001 | -10.06 (-26.78 - 6.66) | 0.2241 | 0.28 (-9.1 - 9.66) | 0.951 |
| Bayley-III Cognitive | ILF-L | 0.4012 | 340 (109 - 571) | 0.005 | 0.389 (0.163 - 0.615) | 0.0013 | -9.48 (-19.13 - 0.18) | 0.0542 | -0.57 (-8.32 - 7.17) | 0.8822 |
|  | Early Infant DTI RD Trajectory |  |  |  |  |  |  |  |  |  |
| 3 Year |  |  |  |  |  |  |  |  |  |  |
| Bayley-III Motor | ILF-R | 0.6452 | -780 (-1511 - -49) | 0.0375 | 0.278 (0.096 - 0.46) | 0.0042 | -12.09 (-18.7 - -5.48) | 0.0009 | 12.97 (7.11 - 18.83) | 0.0001 |
| Bayley-III Language | SLF-R | 0.7268 | -697 (-1307 - -87) | 0.0284 | 0.18 (-0.017 - 0.376) | 0.0695 | -15.23 (-22.99 - -7.47) | 0.0011 | 2.23 (-4.09 - 8.55) | 0.4574 |
|  | Post-Surgical DTI RD Trajectory - No Significant Findings |  |  |  |  |  |  |  |  |  |
|  | Perioperative DTI FA Trajectory |  |  |  |  |  |  |  |  |  |
| 3 Year |  |  |  |  |  |  |  |  |  |  |
| Bayley-III Motor | ILF-L | 0.5365 | -604 (-1135 - -74) | 0.0269 | 0.318 (0.125 - 0.51) | 0.002 | -14.79 (-22.92 - -6.67) | 0.0008 | 12.56 (5.61 - 19.5) | 0.0009 |

Abbreviations: WMV – cerebral white matter volume; DGM – deep grey matter volume; FOF – Fronto-Occipital Fasciculus; ILF – Inferior Longitudinal Fasciculus; SLF – Superior Longitudinal Fasciculus; L – Left; R – Right

### Early Infant Brain Trajectories (Overall Period)and Infant Outcomes

Reduced whole brain volume trajectory (β=0.0008, CI 95%, 0.0001-0.0016; p=0.0286; FC 21.3%) predicted poor 1-year language outcome (Table 2, Figure 2). Within this analysis, low maternal IQ (β=0.2572, CI 95%, 0.0821-0.4323; p=0.0048; FC 36.6%) and presence of genetic abnormality (β=-11.492, CI 95%, -18.8864–-4.0975; p=0.003; FC 40.9%) co-contributed to the poorer 1-year language performance.

### Post-surgical Brain Tractography and Childhood Outcomes

In the post-surgical period imaging trajectories, the only significant findings were between tractography changes and 3-year early childhood cognitive and motor outcomes (Table 2, Figure 3). Reduced FA trajectory of the FOF-R (β=3769.006, CI 95%, 1055.102-6482.911; p=0.0082; FC 22%) predicted – along with lower maternal IQ (β=0.3506, CI 95%, 0.1887-0.5125; p=0.0001; FC 53.6%), presence of genetic abnormality (β=-6.4405, CI 95%, -12.7943--0.0867; p=0.0472; FC 11.7%), and single ventricle status (β=5.8644, CI 95%, 0.2958-11.4330; p=0.0379; FC 12.7%) – poor 3-year cognitive outcome. Reduced FA trajectory of ILF-L (β=2760.332, CI 95%, 201.2291-5319.434; p=0.0354; FC 11.4%) and ILF-R (β=3177.923, CI 95%, 651.1846-5704.661; p=0.0156; FC 18.8%), and an increased RD trajectory of ILF-L (β=-604.336, CI 95%, -1134.79--73.8809; p=0.0269; FC 12.2%) – along with low maternal IQ (β=0.2553, CI 95%, 0.0503-0.4603; p=0.0165, FC up to 27%), presence of genetic abnormality (β=-13.4648, CI 95%, -21.3071--5.6225; p=0.0015, FC up to 35%), and single ventricle status (β=10.6483, CI 95%, 3.6954-17.6012; p=0.004, FC up to 32.3% – predicted poor motor outcome (Figure 3b-c).

**Figure 3.**
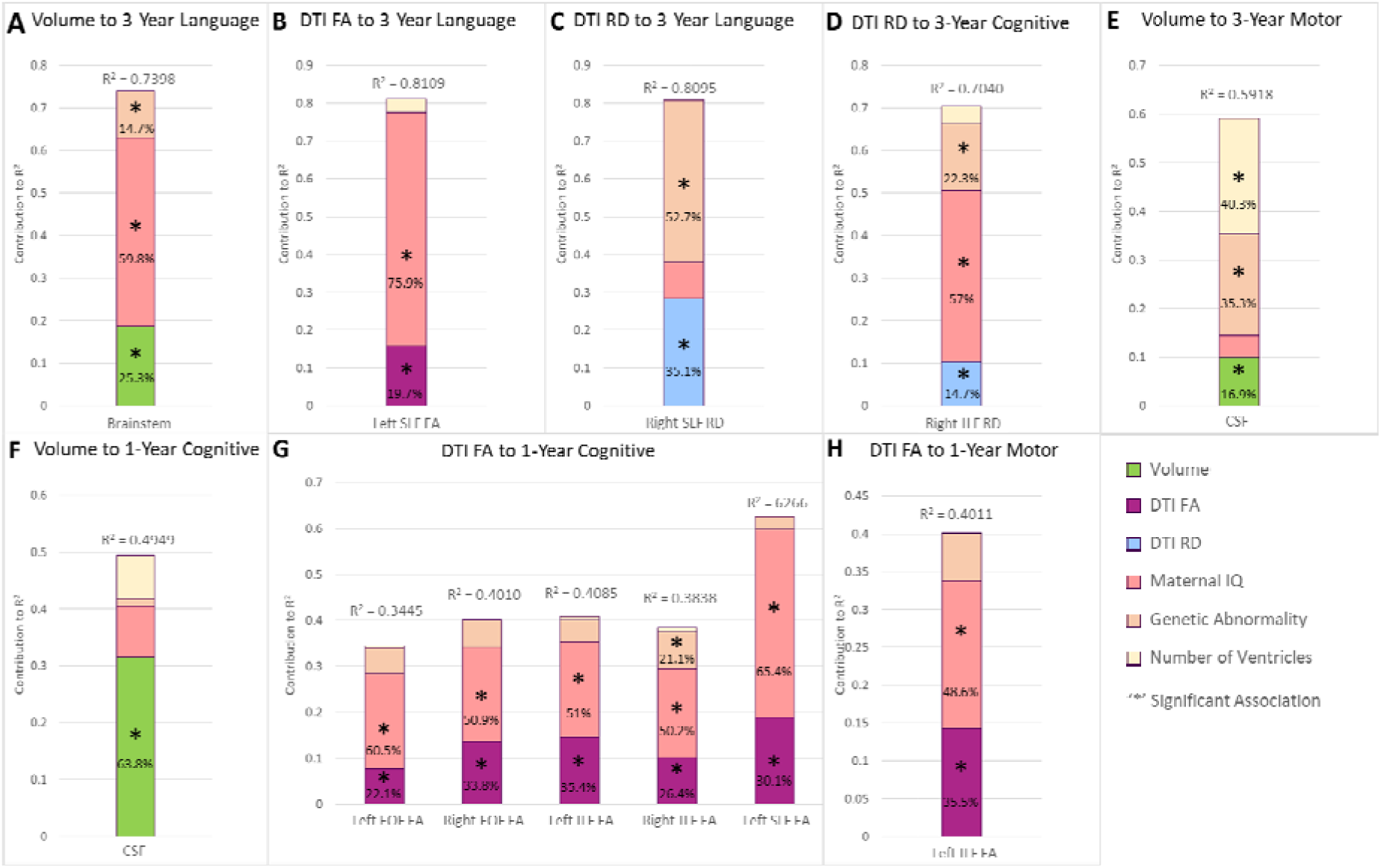
Perioperative period multi-modal imaging trajectories and non-imaging factors contribution to variability in early childhood (3-year outcomes) (A-E) and infant (1-year outcomes) (F-H) outcomes. The peri-operative period imaging trajectories did not predict 5 year outcomes. Macrostructural trajectories (volume) contributed up to 64% in cognitive (cerebral spinal fluid volume increase-F), 17% of motor, and 25% of language variability in 1-year outcomes. Microstructural FA trajectories accounted for up to 30% of cognitive, 36% of motor, and 20% of language variability in 1-year outcomes. Microstructure RD trajectories contributed up to 15% of cognitive and 35% of language outcomes. As such, peri-operative neuroimaging trajectory variables showed remarkable consistency in degree of neurodevelopmental outcome variance explained across 1 year and 3-year time points, with the exception of increased CSF fluid. Genetic abnormality contribution to outcome was relatively consistent between 1-year to 3-year time points, with the exception of language outcomes at 3 years. In contrast, maternal IQ contributed to the outcomes, and the contribution to variability clearly showed a cumulative increase over time from 1-year to 3-year to 3 years old.

### Perioperative Brain Trajectory and Childhood Outcomes

In the perioperative imaging trajectories, reduced trajectory of brainstem volume (β=0.0073, CI 95%, 0.0019-0.0126; p=0.0111; FC 25.3%) as well as reduced FA trajectory of SLF-L (β=154.6626, CI 95%, 46.4283-262.897; p=0.009, FC 19.7%) and increased RD trajectory of SLF-R (β=-66.623, CI 95%, -124.452—8.7941; p=0.0296, FC 35.1%) predicted poor 3-year language performance (Table 2, Figure 4). Along with the imaging findings within these models, low maternal IQ (β=0.5034, CI 95%, 0.3240-0.6827; p<0.0001, FC up to 75.9%) and presence of genetic abnormality (β=-12.3710, CI 95%, -21.1351--3.6069; p=0.0125. FC up to 52.7%) also predicted poor 3-year language performance. Within this imaging epoch, increased RD trajectory of ILF-R (β=-90.6849, CI 95%, -155.0758--26.2940; p=0.0076; FC 14.7%) – along with maternal IQ (β=0.4119, CI 95%, 0.2636-0.5602; p<0.0001, FC 57%) and presence of genetic abnormality (β=-9.6425, CI 95%, -15.1932--4.0918; p=0.0014, FC 22.3%) – predicted poor 3-year cognitive outcome. While increased trajectory of CSF volume (β=-0.0002, CI 95%, -0.0003--0.0001; p=0.0365, FC 16.9%) – along with presence of genetic abnormality (β=- 13.5942, CI 95%, -22.3469--4.8415; p=0.0044, FC 35.3%) and single ventricle status (β=11.7771, CI 95%, 4.6767-18.8775; p=0.0044, FC 40.3%) – predicted poor 3-year motor performance.

### Perioperative Period Brain Trajectory and Infant Outcomes

Increased CSF volume (β=_-_0.0002, CI 95%, -0.0003--0.0001; p=0.0062, FC 63.8%) predicted poorer 1-year cognitive outcome (Table 2, Figure 4). Reduced FA trajectory of FOF-L (β=231.6979, CI 95%, 10.9113-452.4846; p=0.0402, FC 22.1%), FOF-R (β=330.7066, CI 95%, 88.4442-572.9691; p=0.0089, FC 33.8%), ILF-L (β=326.4118, CI 95%, 108.9067-543.9169; p=0.0043, FC 35.4%), ILF-R (β=336.9965, CI 95%, 81.3352-592.6579; p=0.0112, FC 26.4%), and SLF-L (β=256.6343, CI 95%, 90.7343-422.5343; p=0.0042, FC 30.1) predicted poor 1-year cognitive outcomes. Within these findings, lower maternal IQ (β=0.6652, CI 95%, 0.3735-0.9569; p=0.0001, FC up to 65.4%) - and presence of genetic abnormality (β=-10.9218, CI 95%, -20.1926--1.6511; p=0.0222, FC 21.1%) with only the ILF-R finding – also contributed to poor 1-year cognitive outcome.

## Discussion

Prior studies have shown that neurodevelopmental impairments – manifesting as deficits in gross cognitive, visuo-motor, and language functions – are common in children with CHD in infancy and early childhood^15,28,29^. These studies also linked these neurodevelopmental impairments with brain dysmaturation – characterized by MRI biomarkers of qualitative assessments and quantitative volume and WM tractography measures – in the early neonatal period^6,30–32^. However, in these studies, the imaging biomarkers have been modeled as individual exposures on a single time point basis, even in studies that acquired multiple scans. We demonstrated modeling early infant brain trajectories can prognosticate childhood neurodevelopmental outcomes, even after adjusting for important patient-specific, known genetic and socio-environmental factors. The early-infant trajectory encapsulates the microstructural and macrostructural changes the neonatal CHD brain experiences from just after birth to approximately six months of life, which is a major critical period of brain development. We show regions of higher vulnerability include not only white matter structures but also subcortical structures, which is consistent with prior infant CHD neuroimaging literature. ^33–35^

For non-imaging covariates, maternal IQ had the most significant contribution in the multi-variable model, followed by the presence of genetic abnormalities, while number of cardiac ventricles had the least significant predictions. Although maternal IQ has been previously been collected in a CHD related study^36^, our analysis is the first to model the quantitative effects of maternal IQ in this population in conjunction with imaging factors. Parental SES (combined measure of marital and employment status, educational attainment, and occupation prestige) is often used as a proxy for environmental factors and has been shown to influence neurodevelopmental outcomes in other studies.^37–39^ We found that parental SES was highly colinear with maternal IQ, but had significantly fewer correlations with outcomes^40^. Prior research has found that “cognitively stimulating environments”^41^ are associated with development of intelligence and while this is correlated with educational attainment and occupational prestige additional factors such as education quality, parental expectations for academic achievement, and parental scaffolding may explain the unique contribution of maternal IQ above and beyond parent SES^40,42^. Furthermore, higher maternal IQ may be involved in greater parental involvement and further enrichment of a child’s cognitive stimulation. This early stimulation can positively impact CHD neurodevelopmental outcomes as recently demonstrated by a neuroimaging study.^43^ Maternal IQ may also correlate with seeking better care for the child, or even earlier, in seeking prenatal care and making informed decisions during pregnancy. Adequate prenatal care is essential for healthy fetal development and can impact neurodevelopmental outcomes.^44^ This protective and stimulating effect of maternal IQ is supported by our findings where maternal IQ was highly predictive of neurodevelopmental outcomes, and usually provided the highest contribution to the variance (R^2^). Furthermore, the contribution of maternal IQ to outcomes increased in proportion from one to five years, suggesting a cumulative effect on neurodevelopmental over time. In contrast, the impact of genetic abnormalities diminishes during this same period. The presence of genetic abnormality predicted outcomes in all three domains at 1-year and 3-year assessment points, with a significant contribution observed for 3-year language outcomes, but no contribution for 5-year outcomes. Single ventricle status (i.e., patients with HLHS) was limited to almost exclusively predicting poor 3-year motor outcomes, which is consistent with prior literature. Genetic abnormality and single ventricle status are patient-specific conditions present in utero, and the fact that they are not predictive of neurodevelopmental outcomes beyond three years suggests that, by 5-year of age, the gross heritable mechanisms and environmental factors as indexed by maternal IQ might have had time to exert more significant influence on neurodevelopment, and thereby counter the adverse effects of these patient factors.

We also found that modeling earlier brain trajectories identified critical periods of brain development during the peri-operative and post-surgical periods, which predicted earlier ND outcomes (but not later 5-year outcomes). The perioperative period coincides with a period of neonatal neurogenesis that experiences rapid myelination, and perhaps because of this, microstructure trajectories dominate significant findings – with reduced cortical-cortical tracts FA trajectories predicting poor cognitive outcomes at one year. These findings corroborate the WM tract findings from prior studies^7,8,14,45–47^. The post-surgical trajectory showed that reduced white matter microstructure predicted poor cognitive and motor outcomes at three years and may represent correlates of diffuse WMI. Of note, in pre-clinical surgical-based animal models of CHD postnatal subventricular zone is vulnerable to neurotoxicity from volatile anesthetic agents ^48,49^ and hypoxia, resulting in diffuse WMI of white matter tracts, including SLF, ILF, and FOF. Diffuse WMI also correlates with cortical long-range connectivity dysmaturation. In contrast, focal WMI, acquired in CHD infants on serial preoperative/immediate postoperative brain MRIs (usually performed on 7-14 postnatal days and are detected with 3D-T1 based MR imaging), involve punctate periventricular fronto-parietal white matter lesions involving long-range connectivity crossing-fibers,^50–56^ also caused by hypoxia/inflammation. Of note, in our study, focal WMI was not predictive of outcomes in this cohort, and this could be related to the overall incidence of these lesions decreasing over time, as recently described by Peyvandi et al.^57^ Additionally, the influence of positive socio-demographic factors could act as a potential cognitive reserve mechanism, limiting the impact of acquired brain injury patterns with neurocognitive outcomes, which has been noted by Latal et al.^58^

Our study has numerous strengths, including the prospective, longitudinal design, the solid and objective cognitive characterization of the patients and their parents, the racial and ethnic diversity of the sample, and the ability to perform serial scanning within a tight window. However, there are notable weaknesses. We modeled microstructure and macrostructure trajectories as linear changes between time points in this study but acknowledge that this is a simplification necessitated by limitations of imaging resources in the study. Additional scans throughout the early neonatal would provide a more accurate trajectory which may or may not be linear – an approach that should be incorporated into future studies on brain trajectories. Another limiting factor is the relatively high attrition rate, both in terms of lessoning the sample size and potential for introducing bias, in neurodevelopmental testing. However, we did a comparison analysis to ensure no differences in imaging and non-imaging variables between those who were assessed and those who did not participate that would skew or bias the results (see supplement). Finally, unbiased prediction of early childhood neurodevelopmental outcomes requires validated in an independent cohort.

## Conclusion

We have shown that reduced trajectory of multi-modal neonatal brain maturation predicts poorer early childhood outcomes, especially language-based outcomes, despite the contribution of known genetic and demographic factors. Imaging trajectories, with their capability for earlier assessment than neuropsychological tests, and the fact that these imaging trajectories can predict outcomes even up to 5 years, might be helpful as early biomarkers to predict neurodevelopmental outcomes to help guide intervention. Maternal IQ was cumulative over time, exceeding the influence of known innate cardiac and genetic factors in complex CHD, underscoring the importance of heritable and parental-based environmental factors.

## Data Availability

All data produced in the present study are available upon reasonable request to the authors.

## Notes

### Competing Interest Statement

The authors have declared no competing interest.

### Funding Statement

Funding was provided by the National Heart, Lung, and Blood Institute (R01 HL152740-1, R01 HL128818-05).

