## Supplement for "Postnatal Brain Magnetic Resonance Imaging Trajectories and Maternal Intelligence Predict Neurodevelopmental Outcomes in Complex Congenital Heart Disease"

### eMethods

#### MR Imaging Acquisition

Detailed MRI sequence parameters are presented in Supplemental eTable 1.

##### eTable 1: MRI Sequence Parameters

| Image Type | TR/TE (ms) | Flip angle (°) | FOV  (mm) | Slice thickness/ gap (mm) | Number of slices | Acquisition/  reconstruction matrix size | Other |
| --- | --- | --- | --- | --- | --- | --- | --- |
| 3D FFE  T1-weighted sagittal | 20/ 4.1 | 30 | 200 | 1.0/ 0.0 | 100 | 224/256 |  |
| 15 direction  axial DTI | 10,071/90 | 90 | 256 | 2.7/ 0.0 | 55 | 96/128 | b=0,860 (c/mm^2^) |

DTI, Diffusion tensor imaging; DWI, Diffusion weight imaging; FFE, fast field echo; FAIR, fluid-attenuated inversion recovery; FOV, field of view; IR, inversion recovery; PD, proton density; SE, spin echo; SWI, susceptibility-weighted imaging; TE, echo time; TR, relaxation time; TSE, turbo spin echo.

| Image Type | TR/TE (ms) | Flip angle (°) | FOV  (mm) | Slice thickness/ gap (mm) | Number of slices | Acquisition/  reconstruction matrix size | Other |
| --- | --- | --- | --- | --- | --- | --- | --- |
| 3D FFE  T1-weighted sagittal | 20/ 4.1 | 30 | 200 | 1.0/ 0.0 | 100 | 224/256 |  |
| 3D TSE  T2-weighted sagittal | 3200/ 130 | 90 | 200 | 1.0/ 0.0 | 140 | 240/256 |  |
| SE PD/T2-weighted axial | 2500/ 18120 | 90 (refocus angle 140) |  |  |  |  |  |
| FFE axial | 610/ 23 | 15 | 200 | 5.0/ 1.0 | 20 | 256/256 |  |
| TSE T2-weighted coronal FLAIR | 11000/ 140  (TI2600) | 90 | 200 | 3.0/ -1.0 | 53 | 256/256 |  |
| 15 direction  axial DTI | 10,071/90 | 90 | 256 | 2.7/ 0.0 | 55 | 96/128 | b=0,860 (c/mm^2^) |
| Axial DWI | 10,000/ 81  (IR delay 2100) | 90 | 200 | 6.0/ 1.0 | 20 | 112/256 | b=0,1000 (s/mm^2^) |
| 3D SWI axial | 56/40 | 20 | 256 | 2.0/ 0.0- | 32 | 512/512 |  |

#### Regional Brain Volumes

All T1’s for each subject were processed through an in-house semi-automated segmentation pipeline, Neonatal Brain Structure Segmentation (NeBSS)^1^ – an open-source Python project and is freely available at www.github.com/PIRCImagingTools/NeBSS. Additionally, a Docker image and Dockerfile are also available. Subject T1 images are first pre-processed using FSL’s Brain Extraction Tool (BET)^2^ and FSL FAST^3^ for bias correction. After pre-processing, NeBSS has two branches, Branch A which outputs structural volumes using the ALBERT Brain Atlas^4^, and Branch B which outputs probabilistic tissue volumes using the Probabilistic Neonatal Brain Atlas^5^. Within Branch A, four atlas images, chosen based on age similarity to the subject’s gestational age, are transformed into subject space using Advanced Normalization Tools (ANTs) non-linear transformation algorithm^6^. A voxel-wise winner takes all approach is performed and a winner is chosen at random if there is a tie. The resulting segmentation has 50 non-overlapping regions. After completion, inspection and manual correction for the cerebellum, amygdala, and hippocampus is carried out. Branch B works in the opposite direction, mapping the subject into the probabilistic atlas space, again utilizing ANTs non-linear transformation algorithm. Threshold values for each tissue map are manually determined and binarized at that threshold value. Branch B outputs 10 volumes from independent probabilistic tissue maps. The schematic of this image processing pipeline is presented in eFigure 1.

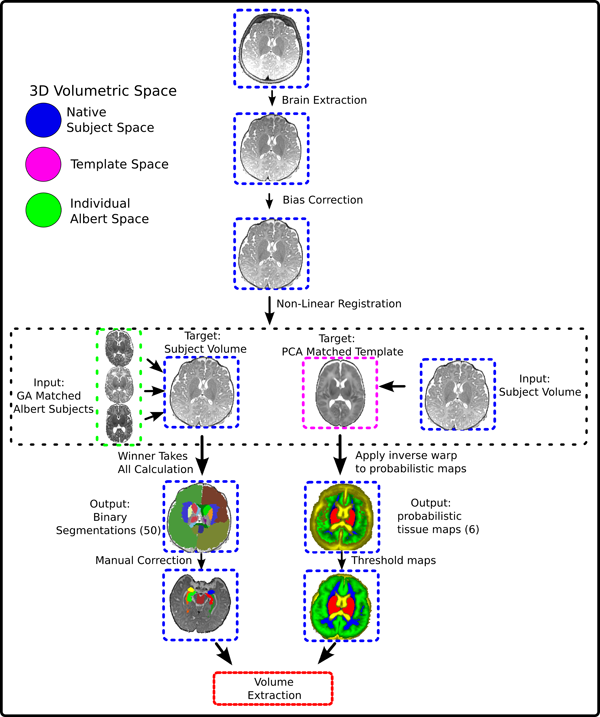

eFigure 1. Diagram showing the Neonatal Brain Structure Segmentation (NeBSS) pipeline used to generate the volume measurements of the various regions. The two parallel processes in the lower half of the diagram illustrates the Winner Takes All (left) and probabilistic maps (right) algorithms used to generate outputs that are ultimately combined to provide extracted volumes of interest.

#### White Matter Tractography

Each DTI acquisition was manually inspected for artifacts, and affected gradient volumes were removed. Each was eddy current and motion corrected with DSI Studio^7^ and reconstructed using the diffusion tensor algorithm. The processed DTI of each participant was used to generate a set of white matter tracks with an in-house automated tractography pipeline which was appropriate for the age-span of this study. A brief explanation of the pipeline is as follows. Fractional anisotropy (FA) maps were generated for each participant from the processed DTI. The participant whose FA map has the lowest deformation – after the process of registering each subject’s FA map to every other subject using FSL^8^ FLIRT^2^ and FNIRT^9^ – is chosen as the most representative subject. This representative subject’s FA map is used as the template space to draw the dedicated sets of regions of interests (ROI) and regions of avoidance (ROA) – which are necessary for generation of individual white matter tracks. These sets of ROI and ROA are then inverse transformed, using ANTS^10^, and back projected onto each participant’s DTI space to generate subject specific set of white matter tracks. Deterministic tractography was conducted in DSI Studio with the following parameters: FA threshold = 0.1, angle threshold = 45°, step size = 1mm, without smoothing, max tracts = 5,000, max seeds = 10,000,000. Example tractography images for these tracts are presented in eFigure 2.

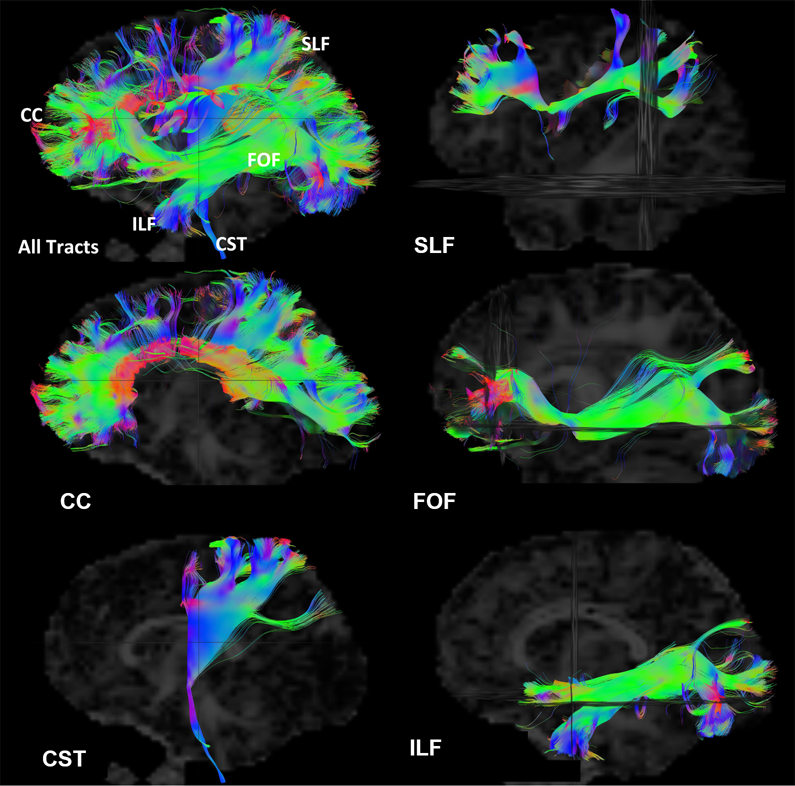

eFigure 2. This figure illustrates the white matter tracts generated with an in-house automated tractography pipeline built upon DSI studio. The corpus callosum (CC) is divided into genu, body, and splenium. The cortico-spinal tract (CST), fronto-occipital fasciculus (FOF), inferior longitudinal fasciculus (ILF), and superior longitudinal fasciculus (SLF) were divided into left and right tracts.

#### WMI segmentation

All MRI series (Scan 1, Scan 2 and Scan 3) underwent review by a pediatric neuroradiologist (J.V.H.) to assess for the presence of brain injury. An “injury” was defined as the presence of any hemorrhagic stroke, ischemic stroke, or white matter injury (lesions were defined as significant if observed injury > 1 mm). For the cases with large enough lesion volume, segmentation was carried out using methods previously described by our group.^11^

#### Neurodevelopmental Tests

The Bayley-III domains tested at 1 and 3, years, and the equivalent conceptual comparison test at 5 years are presented in eTable 2.

##### eTable 2: Tests used in Neurodevelopmental Assessment

| Domain | Included Year 1 Test | Included Year 3 Test | Included Year 5 Test |
| --- | --- | --- | --- |
| **Language** | Bayley-III: Language Composite | Bayley-III: Language Composite | WPPSI-III: Verbal IQ Composite |
| **Cognition** | Bayley-III: Cognitive Composite | Bayley-III: Cognitive Composite | WPPSI-III: Full Scale IQ Composite |
| **Motor** | Bayley-III: Motor Composite | Bayley-III: Motor Composite | Beery VMI: Motor Coordination (+Beery VMI: Visual-Motor Integration*) |

#### Statistical Analysis:

##### Multivariable-model Covariate Selection

The selection of covariates are as follows. The thirteen non-imaging risk factors were examined and include: socio-demographic factors consisting of parental SES, maternal IQ, sex, and race & ethnicity; factors intrinsic to the participant consisting of the presence of genetic abnormalities, 22q deletion, and number of cardiac ventricles; perinatal factors consisting of gestational age at birth and birth weight; medical care and surgery related factors consisting of whether the patient had open sternum surgery, number of days with open sternum, and length of stay at the hospital; and white matter injury. An initial univariable regression analysis was conducted between each risk factor and each of the neurodevelopmental tests. From this initial regression test, the risk factors that demonstrated significant associations (alpha < 0.05) with outcomes are processed through correlation tests for collinearity and then subjected to model selection. This screening multi-variable model selection is composed of non-imaging factors only – without imaging data – and is used as a model reduction to eliminate the predictor variables that are not contributory. The factors that were found to be consistently associated with tests performance within this multi-variable regression screen were incorporated into the final model.

Thus, we investigated the relationship of each imaging trajectory to each neurodevelopmental test using the multi-variable analysis with a fixed set of non-imaging variables as covariates. The results from this model selection process are presented in eResults of this Supplement.

##### Factor Contribution Analysis

After the multi-variable regression analysis, a post-hoc factor contribution analysis was conducted for each model that demonstrated significant (variable specific p<0.05) association between imaging trajectory and neuropsychological test. This post-hoc analysis examined the contributions of each independent variable – imaging, as well as the non-imaging in the final multi-variable model – to the variance in test performance using the formula presented in eEquation 1.

$$Factor Contribution of Variable= \frac{PRE of Variable}{PRE Total}\times R^{2} (eEquation 1)$$

#### Missing NDT Data

There were attritions in the study cohort for neurodevelopmental testing at 1-year, with fewer in the 3-year and 5-year follow up. In order to assess if there were any significant differences in imaging trajectories between those who had NDT and those without NDT (and thus excluded from the main analysis), a comparison analysis using T-test was conducted. The findings are summarized in eResults section of this Supplement.

### eResults

#### Study Design and Cohort of Participants

+++ Need Julia’s Cohort Tracking

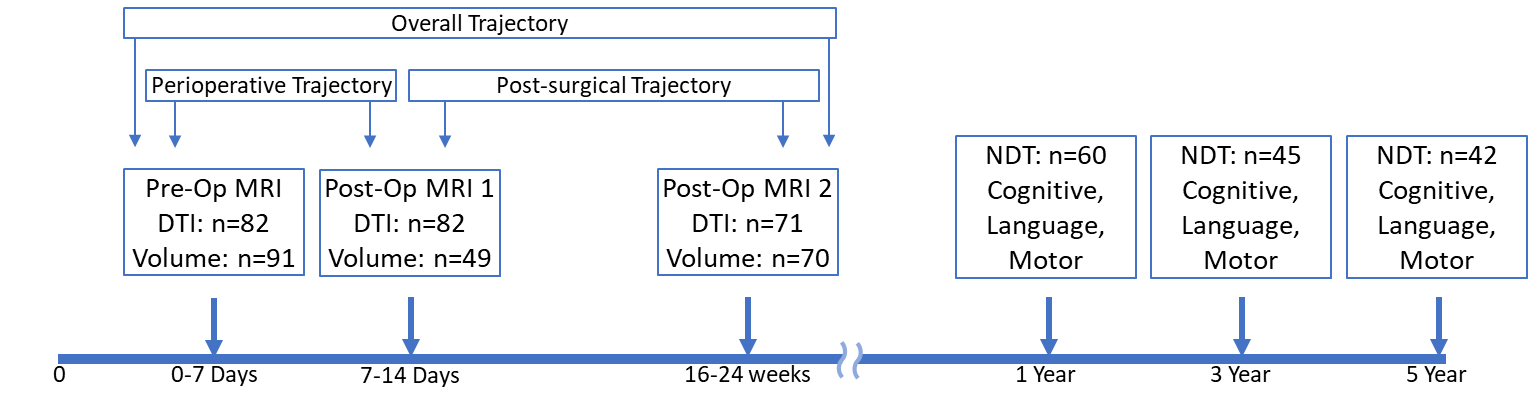

eFigure 1. Overview of imaging and neuropsychological testing time points, along with imaging trajectory scheme. Perioperative Trajectory is between Pre-Op MRI and Post-Op MRI #1. Post-surgical Trajectory is from Post-Op MRI#1 to Post-Op MRI#2. Overall Trajectory encompasses the Pre-Op MRI to Post-Op MRI#2. Neuropsychological tests assessing cognitive, language, and motor were conducted with Bayley-III for one and three years. For five year assessment WPPSI-III Full Scale IQ, WPSSI-III Verbal IQ, and Beery VMI were used for cognitive, language, and motor, respectively.

#### Summary Statistics

The summary statistics of Brain Volumes are presented in eTable 3a. The summary statistics of White Matter Tractography FA and RD measurements are presented in eTable 3b. The summary statistics of Neurodevelopmental Tests Scores are presented in eTable 3c. The summary statistics of Brain Volumes trajectories – given as changes in cubic centimeters over weeks – are presented in eTable 3d. The summary statistics of DTI FA and RD tractography measurements – given as change in FA index and 10^-3^mm2/s/week– are presented in eTable 3e and 3f, respectively.

##### eTable 3a: Volumes Summary Statistics

|  | Pre-Op (N = 91) | Post-Op1 (N = 39) | Post-Op2 (N = 70) |
| --- | --- | --- | --- |
| Volumes (cc) | Mean (SD) | Mean (SD) | Mean (SD) |
| Brainstem | 5289 (674) | 674 (5294) | 5294 (5343) |
| Cerebellum | 24920 (3100) | 3100 (24146) | 24146 (24721) |
| Cortex | 187931 (30171) | 30171 (197105) | 197105 (192434) |
| CSF | 76828 (26167) | 26167 (77355) | 77355 (90893) |
| DGM | 20374 (4588) | 4588 (20594) | 20594 (20320) |
| WM | 150766 (24147) | 24147 (145428) | 145428 (151057) |
| Whole Brain with CSF | 469526 (45068) | 45068 (472092) | 472092 (479494) |
| Whole Brain without CSF | 392698 (43714) | 43714 (392801) | 392801 (388601) |

Abbreviations: DGM - deep grey matter; WM - white matter

##### eTable 3b: Diffusion Tensor Imaging Summary Statistics

|  | Pre-Op (N = 82) | Post-Op1 (N = 82) | Post-Op2 (N = 71) |
| --- | --- | --- | --- |
| Fractional Anisotropy | Mean (SD) | Mean (SD) | Mean (SD) |
| CC Genu | 0.2359 (0.0207) | 0.2371 (0.0215) | 0.3053 (0.0269) |
| CC Body | 0.2117 (0.0194) | 0.208 (0.0211) | 0.265 (0.0242) |
| CC Splenium | 0.257 (0.0235) | 0.2569 (0.0197) | 0.3255 (0.0378) |
| CST-L | 0.2552 (0.0244) | 0.2597 (0.0241) | 0.3502 (0.0323) |
| CST-R | 0.2431 (0.023) | 0.2421 (0.0223) | 0.3407 (0.0368) |
| FOF-L | 0.2057 (0.0187) | 0.209 (0.0185) | 0.2719 (0.0248) |
| FOF-R | 0.2064 (0.0162) | 0.2097 (0.019) | 0.2718 (0.0242) |
| ILF-L | 0.1877 (0.0167) | 0.1938 (0.02) | 0.2662 (0.0244) |
| ILF-R | 0.1963 (0.0172) | 0.1994 (0.0191) | 0.2672 (0.0232) |
| SLF-L | 0.186 (0.0201) | 0.1879 (0.0275) | 0.2394 (0.0246) |
| SLF-R | 0.1787 (0.0237) | 0.1833 (0.0246) | 0.238 (0.023) |
| Radial Diffusivity (10-3mm2/s) |  |  |  |
| CC Genu | 1.2457 (0.0846) | 1.2381 (0.082) | 0.9816 (0.0862) |
| CC bBody | 1.2697 (0.1092) | 1.288 (0.1188) | 1.0589 (0.1106) |
| CC Splenium | 1.2303 (0.11) | 1.2298 (0.0926) | 1.0286 (0.1557) |
| CST-L | 1.0804 (0.0856) | 1.0796 (0.1034) | 0.8782 (0.0837) |
| CST-R | 1.1069 (0.1326) | 1.1223 (0.1154) | 0.8913 (0.086) |
| FOF-L | 1.2188 (0.073) | 1.2051 (0.0805) | 0.9748 (0.0665) |
| FOF-R | 1.212 (0.069) | 1.196 (0.0808) | 0.9487 (0.0624) |
| ILF-L | 1.296 (0.0832) | 1.2963 (0.0978) | 1.0078 (0.1172) |
| ILF-R | 1.2736 (0.0841) | 1.2597 (0.0819) | 0.9679 (0.0713) |
| SLF-L | 1.1615 (0.0911) | 1.1768 (0.1044) | 0.9328 (0.0724) |
| SLF-R | 1.1941 (0.1052) | 1.1823 (0.1055) | 0.95 (0.0979) |

Abbreviations: CC – Corpus Callosum; CST – Cortical Spinal Tract; FOF – Fronto-Occipital Fasciculus; ILF – Inferior Longitudinal Fasciculus L

Fasciculus; SLF – Superior Longitudinal Fasciculus; L – Left; R – Right

##### eTable 3c: Neurodevelopmental Tests Summary Statistics

|  | Mean (SD) |
| --- | --- |
| 5 Year NDT (N=36) |  |
| WPPSI-III Verbal IQ | 94.98 (15.42) |
| WPPSI-III Full Scale IQ | 96.1 (17.7) |
| Beery-VMI Motor | 85.48 (14.25) |
| 3 Year NDT (N=39) |  |
| Bayley-III Language | 95.07 (12.45) |
| Bayley-III Cognitive | 97.07 (10.3) |
| Bayley-III Motor | 98.89 (12.48) |
| 1 Year NDT (N=54) |  |
| Bayley-III Language | 88 (12.56) |
| Bayley-III Cognitive | 101.25 (13.49) |
| Bayley-III Motor | 89.14 (14.19) |

##### eTable 3d: Brain Volume trajectories Summary Statistics

| Units: cc/week | Year 1 | Year 3 | Year 5 |
| --- | --- | --- | --- |
|  | Mean (SD) | Mean (SD) | Mean (SD) |
| Early Infant Trajectory | *(N=54)* | *(N=39)* | *(N=36)* |
| Brainstem | 177 (51) | 183 (52) | 183 (52) |
| Cerebellum | 1571 (482) | 1608 (478) | 1611 (512) |
| Cortex | 8702 (2684) | 8957 (2663) | 8817 (2491) |
| CSF | 3823 (3257) | 4150 (3761) | 4099 (3714) |
| DGM | 867 (384) | 957 (404) | 967 (396) |
| WM | 4761 (1892) | 4810 (1935) | 4842 (1718) |
| Whole Brain with CSF | 18038 (3469) | 18247 (3755) | 18323 (3577) |
| Whole Brain without CSF | 14216 (4015) | 14097 (4277) | 14224 (3832) |
| Post-surgical Trajectory | *(N=50)* | *(N=37)* | *(N=36)* |
| Brainstem | 200 (65) | 213 (58) | 206 (73) |
| Cerebellum | 1850 (515) | 1922 (488) | 1901 (523) |
| Cortex | 9713 (2714) | 9521 (2665) | 9964 (2258) |
| CSF | 4124 (3414) | 4651 (3833) | 4825 (3954) |
| DGM | 933 (398) | 1013 (395) | 1017 (416) |
| WM | 5881 (2315) | 6436 (1795) | 6280 (1855) |
| Whole Brain with CSF | 20433 (3164) | 21093 (2854) | 21203 (3088) |
| Whole Brain without CSF | 16308 (3901) | 16442 (3934) | 16377 (4147) |
| Perioperative Trajectory | *(N=48)* | *(N=36)* | *(N=36)* |
| Brainstem | 93 (567) | -10 (458) | 160 (578) |
| Cerebellum | -92 (2392) | -563 (1862) | -439 (1725) |
| Cortex | 5447 (33022) | 6001 (29748) | 443 (27984) |
| CSF | 5867 (30933) | -3443 (27405) | -1522 (29547) |
| DGM | 608 (3671) | 882 (3623) | 619 (3624) |
| WM | -832 (21705) | -6032 (19698) | -5069 (16936) |
| Whole Brain with CSF | 7823 (22258) | 1152 (20170) | 1103 (20225) |
| Whole Brain without CSF | 1956 (35529) | 4595 (32811) | 2624 (28305) |

##### eTable 3e: DTI FA trajectories Summary Statistics

| Units: FA Index/week | Year 1 | Year3 | Year5 |
| --- | --- | --- | --- |
|  | Mean (SD) | Mean (SD) | Mean (SD) |
| Early Infant Trajectory | *(N=54)* | *(N=39)* | *(N=36)* |
| CCBody | 0.0024 (0.0011) | 0.0024 (0.0012) | 0.0025 (0.0011) |
| CST-L | 0.0047 (0.0015) | 0.0047 (0.0015) | 0.0047 (0.0016) |
| CST-R | 0.0047 (0.0018) | 0.0046 (0.0019) | 0.0046 (0.0019) |
| FOF-L | 0.003 (0.0011) | 0.0029 (0.0011) | 0.003 (0.0011) |
| FOF-R | 0.0031 (0.0012) | 0.0029 (0.0013) | 0.0031 (0.0013) |
| Genu | 0.0034 (0.001) | 0.0034 (0.0011) | 0.0033 (0.0011) |
| ILF-L | 0.0038 (0.0013) | 0.0037 (0.0013) | 0.0038 (0.0014) |
| ILF-R | 0.0036 (0.0013) | 0.0035 (0.0012) | 0.0036 (0.0014) |
| SLF-L | 0.0023 (0.0012) | 0.0024 (0.001) | 0.0022 (0.0011) |
| SLF-R | 0.0029 (0.0016) | 0.003 (0.0019) | 0.0032 (0.0018) |
| Splenium | 0.0031 (0.0014) | 0.0028 (0.0017) | 0.0032 (0.0016) |
| Post-surgical Trajectory | *(N=50)* | *(N=37)* | *(N=36)* |
| CCBody | 0.0028 (0.0011) | 0.003 (0.0011) | 0.003 (0.0011) |
| CST-L | 0.0046 (0.0017) | 0.0045 (0.0017) | 0.0046 (0.0015) |
| CST-R | 0.0051 (0.0021) | 0.0053 (0.0021) | 0.0051 (0.002) |
| FOF-L | 0.0031 (0.0013) | 0.003 (0.0012) | 0.0031 (0.0013) |
| FOF-R | 0.0031 (0.0011) | 0.003 (0.001) | 0.0032 (0.001) |
| Genu | 0.0036 (0.0013) | 0.0037 (0.0012) | 0.0034 (0.0012) |
| ILF-L | 0.0035 (0.0014) | 0.0034 (0.0013) | 0.0033 (0.0014) |
| ILF-R | 0.0036 (0.0014) | 0.0037 (0.0014) | 0.0037 (0.0015) |
| SLF-L | 0.0025 (0.0015) | 0.0028 (0.0014) | 0.0026 (0.0013) |
| SLF-R | 0.0032 (0.0017) | 0.0031 (0.0017) | 0.003 (0.0017) |
| Splenium | 0.0033 (0.0013) | 0.0031 (0.0015) | 0.0033 (0.0015) |
| Perioperative Trajectory | *(N=48)* | *(N=36)* | *(N=36)* |
| CCBody | -0.0033 (0.0169) | -0.003 (0.0147) | -0.0027 (0.0162) |
| CST-L | 0.0035 (0.0231) | 0.003 (0.02) | 0.0027 (0.0212) |
| CST-R | -0.0012 (0.0246) | -0.0036 (0.0236) | -0.0066 (0.023) |
| FOF-L | 0.0032 (0.0176) | 0.0036 (0.0095) | 0.0015 (0.0102) |
| FOF-R | 0.0028 (0.0161) | 0.0018 (0.0081) | 0.0007 (0.008) |
| Genu | 0.0007 (0.0175) | -0.0019 (0.0139) | 0.0011 (0.0166) |
| ILF-L | 0.0061 (0.0165) | 0.0054 (0.0119) | 0.006 (0.0118) |
| ILF-R | 0.0037 (0.015) | 0.0011 (0.0085) | 0.0019 (0.01) |
| SLF-L | 0.0018 (0.0287) | -0.0063 (0.0216) | -0.0039 (0.0215) |
| SLF-R | 0.0011 (0.0233) | 0.0064 (0.0188) | 0.0118 (0.0197) |
| Splenium | 0.0003 (0.0175) | 0.0007 (0.0193) | 0.0002 (0.0175) |

##### eTable 3f: DTI RD trajectories Summary Statistics

| 10^-3^mm/s/week | Year 1 | Year3 | Year5 |
| --- | --- | --- | --- |
|  | Mean (SD) | Mean (SD) | Mean (SD) |
| Early Infant Trajectory | *(N=54)* | *(N=39)* | *(N=36)* |
| CCBody | -0.0102 (0.0056) | -0.0097 (0.0046) | -0.0099 (0.0062) |
| CST-L | -0.0101 (0.0053) | -0.01 (0.006) | -0.0104 (0.0058) |
| CST-R | -0.0109 (0.0102) | -0.0111 (0.0116) | -0.0111 (0.0115) |
| FOF-L | -0.0119 (0.0036) | -0.0119 (0.0036) | -0.0118 (0.0037) |
| FOF-R | -0.0132 (0.0034) | -0.0127 (0.0032) | -0.0124 (0.0034) |
| Genu | -0.0133 (0.0044) | -0.0133 (0.004) | -0.0129 (0.0049) |
| ILF-L | -0.0147 (0.0059) | -0.0147 (0.0063) | -0.0143 (0.0065) |
| ILF-R | -0.0152 (0.0039) | -0.0152 (0.004) | -0.0153 (0.0043) |
| SLF-L | -0.0106 (0.0047) | -0.0095 (0.0047) | -0.0097 (0.0048) |
| SLF-R | -0.0122 (0.0053) | -0.0126 (0.0055) | -0.0133 (0.0049) |
| Splenium | -0.0108 (0.0053) | -0.0098 (0.0051) | -0.0106 (0.0061) |
| Post-surgical Trajectory | *(N=50)* | *(N=37)* | *(N=36)* |
| CCBody | -0.012 (0.0055) | -0.0121 (0.0054) | -0.0113 (0.0056) |
| CST-L | -0.01 (0.0067) | -0.01 (0.0073) | -0.0102 (0.0071) |
| CST-R | -0.0114 (0.0067) | -0.0118 (0.0072) | -0.0117 (0.0074) |
| FOF-L | -0.0117 (0.0043) | -0.0116 (0.0043) | -0.0115 (0.0044) |
| FOF-R | -0.013 (0.0041) | -0.013 (0.0042) | -0.0129 (0.0043) |
| Genu | -0.0135 (0.0048) | -0.0137 (0.005) | -0.0128 (0.0052) |
| ILF-L | -0.0146 (0.0064) | -0.0144 (0.0064) | -0.0136 (0.0062) |
| ILF-R | -0.0152 (0.0045) | -0.016 (0.0044) | -0.0154 (0.0046) |
| SLF-L | -0.0122 (0.0056) | -0.0123 (0.0052) | -0.0106 (0.0042) |
| SLF-R | -0.0121 (0.0073) | -0.0126 (0.0049) | -0.0107 (0.0087) |
| Splenium | -0.0109 (0.0046) | -0.0101 (0.0052) | -0.0097 (0.0053) |
| Perioperative Trajectory | *(N=48)* | *(N=36)* | *(N=36)* |
| CCBody | 0.0183 (0.086) | 0.0209 (0.0386) | 0.0097 (0.0936) |
| CST-L | 0.0021 (0.1104) | 0.01 (0.1299) | -0.0034 (0.1296) |
| CST-R | 0.0096 (0.1345) | 0.0289 (0.1525) | 0.026 (0.1585) |
| FOF-L | -0.0159 (0.0494) | -0.0168 (0.0453) | -0.0145 (0.046) |
| FOF-R | -0.0139 (0.0496) | -0.009 (0.041) | -0.0086 (0.0422) |
| Genu | -0.0025 (0.0402) | 0.0001 (0.0327) | -0.0043 (0.034) |
| ILF-L | 0.0025 (0.0721) | -0.0015 (0.0562) | 0.0102 (0.0695) |
| ILF-R | -0.0133 (0.0595) | -0.0021 (0.0361) | -0.0064 (0.0536) |
| SLF-L | 0.0168 (0.0825) | 0.0473 (0.0783) | 0.0399 (0.0832) |
| SLF-R | 0.0132 (0.095) | -0.0009 (0.0693) | -0.0279 (0.0654) |
| Splenium | 0.002 (0.0944) | 0.0069 (0.0478) | -0.0102 (0.0995) |

#### Results from Non-imaging Factors Selection for Multi-variable Model

Initial regression analysis was conducted between 13 non-imaging risk factors and NDT outcomes, and the results are presented in separate tables as patient soci-demographics (eTable 4a), intrinsic factors (eTable 4b), perinatal factors (eTable 4c), medical care and surgery related factors (eTable 4d), and white matter injury (eTable 4e).Maternal IQ seems the most significant of all the risk factors, with all 9 findings where p < 0.05. Presence of genetic abnormality has the second most significant associations with 8 findings. These two factors are associated with poor performance on cognitive, language, and motor tests, and consistent throughout all three NDT epochs. Parental SES showed significant correlation with cognitive, language, and motor in infancy, and cognitive and language in 3-year early childhood NDT. SES seems to be less of a factor in a child's performance as they age, as there were no correlations to 5-Year early childhood NDT. Length of hospital stay and ethnicity were found to correlate with infant NDT outcomes but were largely not correlated with early childhood assessment. Number of cardiac ventricles and sex had one significant correlation. Birthweight, along with whether the participant had open sternum and the duration of the open sternum had no correlations.

To test whether these environmental and clinical risk factors were independent or whether there were overlaps between them, a correlation analysis among these non-imaging variables was conducted (eTable 4f) to test for collinearity. Maternal IQ and Parental SES had a significantly high correlation (Pearson’s R = 0.6798) indicating that these two factors were colinear. Parental SES and Maternal IQ were also correlated with ethnicity and race, indicating multicollinearities between these socio-demographic factors.

After further examination of the ethnicity and race data revealed that this variable was not clearly separable since the attrition of Black and Asian participants in the 3- and 5-year NDT leading to two and zero participants, respectively. This observation and the collinearity of ethnicity with SES, as well as the fact that Ethnicity and race had the least significant findings in the univariable regression with NDT (eTable 4a) lead to its removal from consideration from the multi-variable model.

##### eTable 4a: Comparison of Neurodevelopmental Outcomes to Non-imaging Factors – Socio-Demographic

|  | Parental SES |  | Maternal IQ |  |
| --- | --- | --- | --- | --- |
|  | β Coefficient (95% CI) | p-value | β Coefficient (95% CI) | p-value |
| 5 Year NDT |  |  |  |  |
| WPPSI-III Verbal IQ | 0.352 (-0.015 - 0.719) | 0.0595 | 0.609 (0.32 - 0.897) | **0.0001** |
| WPPSI-III Full Scale IQ | 0.245 (-0.19 - 0.68) | 0.2606 | 0.679 (0.342 - 1.016) | **0.0002** |
| Beery-VMI Motor | 0.094 (-0.218 - 0.406) | 0.5466 | 0.35 (0.105 - 0.596) | **0.0064** |
| 3 Year NDT |  |  |  |  |
| Bayley-III Language | 0.492 (0.265 - 0.719) | **<.0001** | 0.414 (0.226 - 0.601) | **<.0001** |
| Bayley-III Cognitive | 0.271 (0.059 - 0.482) | **0.0134** | 0.332 (0.176 - 0.489) | **0.0001** |
| Bayley-III Motor | 0.158 (-0.11 - 0.426) | 0.2407 | 0.242 (0.028 - 0.457) | **0.0277** |
| 1 Year NDT |  |  |  |  |
| Bayley-III Language | 0.339 (0.109 - 0.569) | **0.0047** | 0.276 (0.091 - 0.461) | **0.0042** |
| Bayley-III Cognitive | 0.324 (0.086 - 0.561) | **0.0086** | 0.302 (0.102 - 0.503) | **0.0039** |
| Bayley-III Motor | 0.261 (0.009 - 0.514) | **0.0427** | 0.275 (0.063 - 0.486) | **0.0118** |
|  | Sex |  | Ethnicity |  |
|  | β Coefficient (95% CI) | p-value | β Coefficient (95% CI) | p-value |
| 5 Year NDT |  |  |  |  |
| WPPSI-III Verbal IQ | -8.071 (-17.824 - 1.681) | 0.1021 | 7.226 (-4.562 - 19.013) | 0.2224 |
| WPPSI-III Full Scale IQ | -8.064 (-19.505 - 3.378) | 0.1618 | 5.738 (-8.036 - 19.513) | 0.4043 |
| Beery-VMI Motor | -1.514 (-10.592 - 7.564) | 0.7379 | 2.65 (-7.273 - 12.573) | 0.5924 |
| 3 Year NDT |  |  |  |  |
| Bayley-III Language | -5.728 (-13.442 - 1.985) | 0.1415 | 5.503 (-3.146 - 14.152) | 0.2063 |
| Bayley-III Cognitive | -4.662 (-11.045 - 1.722) | 0.1481 | 6.345 (-0.677 - 13.366) | 0.0754 |
| Bayley-III Motor | -7.586 (-15.161 - -0.011) | **0.0497** | 5.027 (-3.665 - 13.719) | 0.2499 |
| 1 Year NDT |  |  |  |  |
| Bayley-III Language | 1.319 (-5.357 - 7.995) | 0.6939 | 9.673 (2.974 - 16.373) | **0.0054** |
| Bayley-III Cognitive | 1.286 (-5.836 - 8.408) | 0.7191 | 9.722 (2.49 - 16.955) | **0.0093** |
| Bayley-III Motor | 1.5 (-6.039 - 9.039) | 0.6918 | 13.786 (6.555 - 21.016) | **0.0003** |

##### eTable 4b: Comparison of Neurodevelopmental Outcomes to Non-imaging Intrinsic Factors

|  | Presence of Genetic Abnormalities |  | Genetics: 22q deletion |  | Number of Cardiac Ventricles |  |
| --- | --- | --- | --- | --- | --- | --- |
|  | β Coefficient (95% CI) | p-value | β Coefficient (95% CI) | p-value | β Coefficient (95% CI) | p-value |
| 5 Year NDT |  |  |  |  |  |  |
| WPPSI-III Verbal IQ | 12.823 (2.009 - 23.636) | **0.0213** | -4.583 (-21.226 - 12.06) | 0.5431 | 2.48 (-7.584 - 12.545) | 0.6209 |
| WPPSI-III Full Scale IQ | 15.344 (2.387 - 28.302) | **0.0215** | -5.1 (-28.198 - 17.998) | 0.6177 | 1.292 (-10.551 - 13.135) | 0.8264 |
| Beery-VMI Motor | 13.051 (2.91 - 23.191) | **0.013** | 6.167 (-24.165 - 36.498) | 0.6454 | -1.028 (-10.113 - 8.057) | 0.8203 |
| 3 Year NDT |  |  |  |  |  |  |
| Bayley-III Language | 9.636 (1.601 - 17.672) | **0.0199** | 3.333 (-13.881 - 20.548) | 0.6753 | 0.679 (-6.91 - 8.267) | 0.8577 |
| Bayley-III Cognitive | 8.159 (1.536 - 14.782) | **0.0169** | -2.833 (-11.937 - 6.271) | 0.5038 | -3.429 (-9.617 - 2.76) | 0.2701 |
| Bayley-III Motor | 10.985 (3.099 - 18.871) | **0.0074** | -0.667 (-13.938 - 12.605) | 0.9131 | -7.917 (-15.121 - -0.712) | **0.032** |
| 1 Year NDT |  |  |  |  |  |  |
| Bayley-III Language | 8.892 (1.031 - 16.752) | **0.0273** | 5.167 (-11.452 - 21.785) | 0.5042 | -0.751 (-7.379 - 5.876) | 0.8213 |
| Bayley-III Cognitive | 7.813 (-0.732 - 16.357) | 0.0724 | 0 (-23.012 - 23.012) | 1 | -4.688 (-11.625 - 2.25) | 0.1815 |
| Bayley-III Motor | 10.213 (1.349 - 19.076) | **0.0247** | -1 (-20.154 - 18.154) | 0.9097 | -2.299 (-9.762 - 5.165) | 0.5399 |

Abbreviations: Yr5 – 5-Year Neurodevelopmental Testing Outcome; Yr3 – 3-Year Neurodevelopmental Testing Outcome; Yr1 – 1-Year Neurodevelopmental Testing Outcome.

##### eTable 4c: Comparison of Neurodevelopmental Outcomes to Non-imaging Perinatal Factors

|  | Gestational Age at Birth |  | Birth Weight |  |
| --- | --- | --- | --- | --- |
|  | β Coefficient (95% CI) | p-value | β Coefficient (95% CI) | p-value |
| 5 Year NDT |  |  |  |  |
| WPPSI-III Verbal IQ | -1.158 (-4.724 - 2.409) | 0.5154 | 0.004 (-0.007 - 0.014) | 0.4641 |
| WPPSI-III Full Scale IQ | -1.081 (-5.208 - 3.046) | 0.5991 | 0.005 (-0.007 - 0.017) | 0.3936 |
| Beery-VMI Motor | -1.284 (-4.615 - 2.046) | 0.4404 | 0.001 (-0.008 - 0.011) | 0.7664 |
| 3 Year NDT |  |  |  |  |
| Bayley-III Language | -0.111 (-3.325 - 3.104) | 0.9448 | 0 (-0.009 - 0.008) | 0.9562 |
| Bayley-III Cognitive | 0.75 (-1.907 - 3.408) | 0.5719 | 0.004 (-0.003 - 0.011) | 0.215 |
| Bayley-III Motor | 0.355 (-2.892 - 3.602) | 0.8264 | 0.005 (-0.003 - 0.014) | 0.2061 |
| 1 Year NDT |  |  |  |  |
| Bayley-III Language | 0.779 (-1.946 - 3.504) | 0.5693 | 0.001 (-0.005 - 0.008) | 0.6608 |
| Bayley-III Cognitive | 0.206 (-2.72 - 3.132) | 0.8883 | -0.001 (-0.008 - 0.006) | 0.767 |
| Bayley-III Motor | 0.564 (-2.519 - 3.647) | 0.7156 | -0.001 (-0.008 - 0.007) | 0.8312 |

Abbreviations: Yr5 – 5-Year Neurodevelopmental Testing Outcome; Yr3 – 3-Year Neurodevelopmental Testing Outcome; Yr1 – 1-Year Neurodevelopmental Testing Outcome.

##### eTable 4d: Comparison of Neurodevelopmental Outcomes to Medical Care & Surgery Factors

|  | Hospital Length of Stay (days) |  | Had Open Sternum |  | Number of Days with Open Sternum |  |
| --- | --- | --- | --- | --- | --- | --- |
|  | β Coefficient (95% CI) | p-value | β Coefficient (95% CI) | p-value | β Coefficient (95% CI) | p-value |
| 5 Year NDT |  |  |  |  |  |  |
| WPPSI-III Verbal IQ | -0.096 (-0.365 - 0.173) | 0.4756 | -3.038 (-16.221 - 10.145) | 0.6437 | 5.962 (-8.836 - 20.759) | 0.3479 |
| WPPSI-III Full Scale IQ | -0.08 (-0.446 - 0.285) | 0.6593 | 0.069 (-16.19 - 16.327) | 0.9932 | 6.7 (-13.145 - 26.545) | 0.4016 |
| Beery-VMI Motor | -0.153 (-0.394 - 0.088) | 0.2068 | 7.691 (-3.499 - 18.881) | 0.1725 | 1.484 (-25.074 - 28.042) | 0.8957 |
| 3 Year NDT |  |  |  |  |  |  |
| Bayley-III Language | -0.106 (-0.222 - 0.01) | 0.0725 | 2.529 (-6.548 - 11.605) | 0.5772 | 8.25 (-2.611 - 19.111) | 0.1179 |
| Bayley-III Cognitive | -0.094 (-0.189 - 0.001) | 0.0524 | 3.3 (-4.165 - 10.765) | 0.3776 | 4.286 (-6.716 - 15.287) | 0.3952 |
| Bayley-III Motor | -0.131 (-0.245 - -0.017) | **0.0248** | 4.871 (-4.131 - 13.873) | 0.2812 | 5.036 (-8.285 - 18.356) | 0.4087 |
| 1 Year NDT |  |  |  |  |  |  |
| Bayley-III Language | -0.116 (-0.224 - -0.008) | **0.0354** | -0.602 (-9.405 - 8.201) | 0.8916 | 4.821 (-5.811 - 15.453) | 0.3262 |
| Bayley-III Cognitive | -0.164 (-0.276 - -0.052) | **0.0049** | -2.1 (-11.516 - 7.316) | 0.6569 | 4.643 (-12.198 - 21.483) | 0.5427 |
| Bayley-III Motor | -0.158 (-0.278 - -0.039) | **0.0104** | 5.702 (-4.126 - 15.53) | 0.2502 | -1.286 (-20.304 - 17.732) | 0.88 |

##### eTable 4e: Comparison of Neurodevelopmental Outcomes to Non-imaging Factors – White Matter Injury

|  | WMI |  |
| --- | --- | --- |
|  | β Coefficient (95% CI) | p-value |
| 5 Year NDT |  |  |
| WPPSI-III Verbal IQ | 7.033 (-2.081 - 16.146) | 0.1265 |
| WPPSI-III Full Scale IQ | 6.839 (-3.992 - 17.671) | 0.2087 |
| Beery-VMI Motor | 4.095 (-4.661 - 12.852) | 0.35 |
| 3 Year NDT |  |  |
| Bayley-III Language | 3.323 (-3.252 - 9.898) | 0.3136 |
| Bayley-III Cognitive | 1.963 (-4.219 - 8.145) | 0.5252 |
| Bayley-III Motor | 2.161 (-5.419 - 9.742) | 0.5681 |
| 1 Year NDT |  |  |
| Bayley-III Language | 2.004 (-4.607 - 8.614) | 0.5462 |
| Bayley-III Cognitive | 1.561 (-5.577 - 8.699) | 0.6631 |
| Bayley-III Motor | 2.951 (-4.616 - 10.518) | 0.4379 |

##### eTable 4f: Correlation Among Non-Imaging Factors

|  |  | **Maternal IQ** | **Cardiac Ventricles** | **Sex** | **Birth weight** | **Hosp. Length of Stay** | **ethnicity** | **Race** | **Gen. Abn.** |
| --- | --- | --- | --- | --- | --- | --- | --- | --- | --- |
| **Parental SES** | Pearson R | 0.6798 | 0.0287 | 0.1793 | -0.0522 | -0.2205 | -0.3057 | 0.1957 | 0.1477 |
|  | p-value | **<.0001** | 0.8291 | 0.1743 | 0.6945 | 0.0934 | **0.0185** | 0.1375 | 0.2643 |
| **Maternal IQ** | Pearson R |  | -0.0426 | 0.0871 | 0.0561 | -0.1294 | -0.4928 | 0.29 | 0.109 |
|  | p-value |  | 0.7339 | 0.4866 | 0.6545 | 0.3005 | **<.0001** | **0.0182** | 0.3835 |
| **Cardiac Ventricles** | Pearson R |  |  | -0.0705 | 0.1912 | -0.3768 | 0.0009 | -0.0114 | 0.1474 |
|  | p-value |  |  | 0.497 | 0.0649 | **0.0002** | 0.9929 | 0.9129 | 0.1539 |
| **Sex** | Pearson R |  |  |  | 0.1514 | -0.0034 | -0.0994 | -0.0544 | -0.0848 |
|  | p-value |  |  |  | 0.1408 | 0.9738 | 0.3329 | 0.5964 | 0.414 |
| **Birth weight** | Pearson R |  |  |  |  | -0.1408 | -0.0639 | 0.0784 | -0.0451 |
|  | p-value |  |  |  |  | 0.1713 | 0.5365 | 0.4476 | 0.6661 |
| **Hosp stay** | Pearson R |  |  |  |  |  | 0.2459 | -0.0005 | 0.2052 |
|  | p-value |  |  |  |  |  | **0.0152** | 0.9961 | **0.046** |
| **ethnicity** | Pearson R |  |  |  |  |  |  | 0.2612 | 0.0221 |
|  | p-value |  |  |  |  |  |  | **0.0098** | 0.8316 |
| **race** | Pearson R |  |  |  |  |  |  |  | 0.1799 |
|  | p-value |  |  |  |  |  |  |  | 0.0811 |

#### Multivariable-Model for Adjusted Regression

Based on the findings thus far (eTables 4a-f), it was determined that Maternal IQ, Parental SES, presence of genetic abnormality, and length of hospital stay maybe significant contributors to NDT variance, and they were considered as possible covariates for inclusion in the multi-variable model comparing imaging trajectories to NDT outcomes. While number of cardiac ventricles did not show high level of correlation to NDT on its own, this factor modeled for the nature of CHD lesion and was also considered for inclusion. Consequently, two competing multi-variable regression analyses were run to determine whether to include Maternal IQ or Parental SES. Due to the high collinearity between Maternal IQ and Parental SES, it was deemed the two factors are not independent and including both in the same model would be redundant. Each multivariable model had Maternal IQ or Parental SES and included the other factors of interest as covariates – number of cardiac ventricles, length of hospital stay, and presence of genetic abnormality.

The results from these two competing analyses (eTables 5a and 5b) clearly demonstrate that in the multi-variable model, in the presence of all the other non-imaging factors, Maternal IQ retained significant associations with most of the neurocognitive tests, while Parental SES was only associated with two of the neurocognitive tests. Length of Hospital Stay was also found to not be associated in the presence of other factors. Based on this examination, Maternal IQ was chosen for the final multivariate model, and length of hospital stay and Parental SES were not included.

##### eTable 5a: Maternal IQ Multi-variable Regression

|  | Maternal IQ | | Number of Cardiac Ventricles | | Presence of Genetic Abnormalities | | Hospital Length of Stay (days) | |
| --- | --- | --- | --- | --- | --- | --- | --- | --- |
|  | β Coefficient (95% CI) | p-value | β Coefficient (95% CI) | p-value | β Coefficient (95% CI) | p-value | β Coefficient (95% CI) | p-value |
| 5 Year NDT |  |  |  |  |  |  |  |  |
| WPPSI-III Verbal IQ | 0.571 (0.281 - 0.861) | **0.0003** | -1.166 (-10.708 - 8.376) | 0.8054 | 11.044 (0.803 - 21.285) | **0.0353** | 0.024 (-0.222 - 0.27) | 0.8425 |
| WPPSI-III Full Scale IQ | 0.65 (0.31 - 0.99) | **0.0005** | -4.483 (-15.609 - 6.642) | 0.4182 | 14.005 (1.705 - 26.305) | **0.0269** | 0.082 (-0.24 - 0.404) | 0.6087 |
| Beery-VMI Motor | 0.331 (0.096 - 0.566) | **0.0071** | -3.053 (-12.231 - 6.126) | 0.504 | 12.49 (2.056 - 22.924) | **0.0204** | -0.044 (-0.285 - 0.197) | 0.7115 |
| 3 Year NDT |  |  |  |  |  |  |  |  |
| Bayley-III Language | 0.413 (0.239 - 0.587) | **<.0001** | -0.826 (-7.309 - 5.656) | 0.798 | 8.958 (2.061 - 15.855) | **0.0122** | -0.064 (-0.167 - 0.04) | 0.2204 |
| Bayley-III Cognitive | 0.352 (0.216 - 0.489) | **<.0001** | -5.526 (-10.614 - -0.438) | **0.034** | 8.882 (3.469 - 14.295) | **0.0019** | -0.027 (-0.107 - 0.054) | 0.5117 |
| Bayley-III Motor | 0.28 (0.104 - 0.456) | **0.0026** | -10.231 (-16.8 - -3.663) | **0.0031** | 12.532 (5.543 - 19.52) | **0.0008** | -0.027 (-0.131 - 0.078) | 0.6056 |
| 1 Year NDT |  |  |  |  |  |  |  |  |
| Bayley-III Language | 0.284 (0.105 - 0.463) | **0.0025** | -0.476 (-7.332 - 6.379) | 0.8896 | 9.594 (1.702 - 17.486) | **0.0182** | -0.065 (-0.181 - 0.05) | 0.263 |
| Bayley-III Cognitive | 0.309 (0.117 - 0.501) | **0.0022** | -3.803 (-11.159 - 3.552) | 0.3042 | 9.31 (0.842 - 17.778) | **0.0318** | -0.084 (-0.208 - 0.04) | 0.1797 |
| Bayley-III Motor | 0.276 (0.075 - 0.477) | **0.008** | -1.031 (-8.732 - 6.669) | 0.7892 | 10.49 (1.624 - 19.355) | **0.0213** | -0.101 (-0.231 - 0.028) | 0.1224 |

##### eTable 5b: Parental SES Multivariable Regression

|  | Parental SES | | Number of Cardiac Ventricles | | Presence of Genetic Abnormalities | | Hospital Length of Stay (days) | |
| --- | --- | --- | --- | --- | --- | --- | --- | --- |
|  | β Coefficient (95% CI) | p-value | β Coefficient (95% CI) | p-value | β Coefficient (95% CI) | p-value | β Coefficient (95% CI) | p-value |
| 5 Year NDT |  |  |  |  |  |  |  |  |
| WPPSI-III Verbal IQ | 0.326 (-0.031 - 0.683) | 0.0722 | -0.065 (-10.752 - 10.622) | 0.9903 | 13.341 (1.842 - 24.839) | **0.0243** | -0.018 (-0.294 - 0.257) | 0.8928 |
| WPPSI-III Full Scale IQ | 0.231 (-0.19 - 0.652) | 0.2729 | -2.193 (-14.889 - 10.502) | 0.7274 | 17.136 (3.128 - 31.143) | **0.018** | -0.028 (-0.39 - 0.335) | 0.8778 |
| Beery-VMI Motor | 0.053 (-0.236 - 0.342) | 0.7134 | 1.596 (-7.591 - 10.784) | 0.7262 | 13.173 (3.009 - 23.338) | **0.0126** | -0.116 (-0.353 - 0.121) | 0.3257 |
| 3 Year NDT |  |  |  |  |  |  |  |  |
| Bayley-III Language | 0.463 (0.243 - 0.683) | **0.0001** | -1.85 (-8.986 - 5.286) | 0.6025 | 9.241 (1.369 - 17.113) | **0.0226** | -0.03 (-0.143 - 0.084) | 0.6004 |
| Bayley-III Cognitive | 0.249 (0.045 - 0.454) | **0.0184** | -4.053 (-10.687 - 2.581) | 0.2235 | 7.212 (-0.106 - 14.53) | 0.0532 | -0.035 (-0.14 - 0.071) | 0.5099 |
| Bayley-III Motor | 0.142 (-0.091 - 0.375) | 0.2246 | -10.389 (-17.943 - -2.835) | **0.0084** | 11.534 (3.201 - 19.867) | **0.008** | -0.027 (-0.147 - 0.093) | 0.6491 |
| 1 Year NDT |  |  |  |  |  |  |  |  |
| Bayley-III Language | 0.302 (0.064 - 0.539) | **0.014** | -1.17 (-8.514 - 6.174) | 0.7499 | 5.632 (-2.934 - 14.198) | 0.1923 | -0.074 (-0.195 - 0.048) | 0.2311 |
| Bayley-III Cognitive | 0.275 (0.036 - 0.514) | **0.0248** | -2.647 (-10.025 - 4.731) | 0.4738 | 4.852 (-3.754 - 13.458) | 0.2623 | -0.102 (-0.224 - 0.021) | 0.1012 |
| Bayley-III Motor | 0.192 (-0.056 - 0.441) | 0.1265 | -0.34 (-8.027 - 7.348) | 0.9295 | 6.907 (-2.059 - 15.874) | 0.1279 | -0.127 (-0.255 - 0.001) | 0.0511 |

#### White Matter Injury Results

There were 46 cases identified with white matter injury. There were 35 cases with WMI in the Pre-Op imaging, 7 of which had large enough lesions for segmentation and lesion volume measurement (of these only 4 had NDT). In the Post-Op1 scans 11 new cases of WMI developed while 1 of the prior cases resolved, for a total of 45 cases with post-op WMI. Of these, 17 of the cases had large enough measurable lesion volume (of these 15 had NDT).

Comparison analysis between participants with and without WMI for differences in volume trajectories, DTI FA and RD trajectories, and NDT are presented in eTable 6a, 6b, 6c, and 6d, respectively. There were no significant findings of consequence. Despite the low sample size, a regression analysis between white matter lesion volumes (both pre-op and post-op) and NDT outcomes (against each of the three testing time points) were conducted, but there were no significant findings.

##### eTable 6a: VOLUME Trajectory differences between the groups with and without WMI

|  | Without WMI  (N=16) | With WMI (N=26) |  |  |
| --- | --- | --- | --- | --- |
| Volumes (cc/week) | Mean (SD) | Mean (SD) | t-value | p-value |
| Early Infant Trajectory |  |  |  |  |
| Brainstem | 171 (51) | 182 (58) | -0.79 | 0.4327 |
| Cerebellum | 1430 (498) | 1627 (475) | -1.61 | 0.1112 |
| Cortex | 7707 (3049) | 8711 (3000) | -1.31 | 0.1941 |
| CSF | 3600 (3899) | 4252 (4704) | -0.58 | 0.5645 |
| DGM | 817 (467) | 892 (411) | -0.69 | 0.4919 |
| WM | 4343 (1856) | 4978 (2382) | -1.13 | 0.2624 |
| Whole Brain with CSF | 17275 (3496) | 18259 (3496) | -1.11 | 0.2716 |
| Whole Brain without CSF | 13676 (4538) | 14007 (5015) | -0.27 | 0.7889 |
| Post-Surgical Trajectory |  |  |  |  |
| Brainstem | 199 (49) | 202 (71) | -0.13 | 0.901 |
| Cerebellum | 1659 (639) | 1819 (537) | -0.74 | 0.4642 |
| Cortex | 7874 (3912) | 9464 (2867) | -1.31 | 0.2 |
| CSF | 3697 (3658) | 4088 (4343) | -0.25 | 0.8055 |
| DGM | 790 (391) | 1023 (404) | -1.54 | 0.1345 |
| WM | 5187 (2687) | 5590 (2509) | -0.42 | 0.6806 |
| Whole Brain with CSF | 18807 (4348) | 20549 (3195) | -1.29 | 0.207 |
| Whole Brain without CSF | 15110 (5422) | 16461 (4379) | -0.76 | 0.4542 |
| Perioperative Trajectory |  |  |  |  |
| Brainstem | -50 (680) | 170 (480) | -1.15 | 0.2586 |
| Cerebellum | 510 (1804) | 25 (2626) | 0.58 | 0.5675 |
| Cortex | 10441 (22473) | 5883 (34312) | 0.42 | 0.6778 |
| CSF | -9228 (34261) | 8392 (28359) | -1.67 | 0.1042 |
| DGM | 1610 (3983) | -322 (4148) | 1.35 | 0.1852 |
| WM | -3263 (21229) | 78 (23967) | -0.41 | 0.6819 |
| Whole Brain with CSF | 10844 (19890) | 6654 (23103) | 0.54 | 0.5915 |
| Whole Brain without CSF | 20072 (38090) | -1738 (27808) | 2 | 0.0535 |

##### eTable 6b: DTI FA Trajectory differences between the groups with and without WMI

|  | Without WMI  (N=16) | With WMI  (N=42) |  |  |
| --- | --- | --- | --- | --- |
| Fractional Anisotropy Trajectories (FA Index / week) | Mean (SD) | Mean (SD) | t-value | p-value |
| Early Infant Trajectory |  |  |  |  |
| CC Genu | 0.0031 (0.0014) | 0.0036 (0.0012) | -1.53 | 0.1306 |
| CC Body | 0.002 (0.0009) | 0.0027 (0.0011) | -2.72 | **0.0086** |
| CC Splenium | 0.0031 (0.0018) | 0.0029 (0.0016) | 0.36 | 0.7233 |
| CST-L | 0.0047 (0.0016) | 0.0046 (0.0015) | 0.21 | 0.8317 |
| CST-R | 0.0049 (0.002) | 0.0047 (0.0019) | 0.45 | 0.6563 |
| FOF-L | 0.0033 (0.0012) | 0.0031 (0.0012) | 0.52 | 0.6077 |
| FOF-R | 0.0031 (0.0011) | 0.0032 (0.0015) | -0.24 | 0.8126 |
| ILF-L | 0.0038 (0.001) | 0.004 (0.0014) | -0.5 | 0.6187 |
| ILF-R | 0.0033 (0.0012) | 0.0037 (0.0016) | -0.94 | 0.3525 |
| SLF-L | 0.0021 (0.0014) | 0.0025 (0.0013) | -1.01 | 0.3178 |
| SLF-R | 0.0027 (0.0011) | 0.0031 (0.0021) | -0.8 | 0.4308 |
| Post-Surgical Trajectory |  |  |  |  |
| CC Genu | 0.0032 (0.0013) | 0.0037 (0.0014) | -1.53 | 0.131 |
| CC Body | 0.0025 (0.0009) | 0.0033 (0.0013) | -2.93 | **0.0049** |
| CC Splenium | 0.0031 (0.0016) | 0.0034 (0.0016) | -0.79 | 0.4349 |
| CST-L | 0.0045 (0.0019) | 0.0047 (0.0014) | -0.42 | 0.6744 |
| CST-R | 0.005 (0.0023) | 0.0057 (0.0022) | -1.03 | 0.3104 |
| FOF-L | 0.0031 (0.0013) | 0.0033 (0.0015) | -0.55 | 0.5852 |
| FOF-R | 0.0033 (0.0012) | 0.0033 (0.0012) | -0.15 | 0.8841 |
| ILF-L | 0.0036 (0.0014) | 0.0039 (0.0015) | -0.64 | 0.5256 |
| ILF-R | 0.0034 (0.0012) | 0.0039 (0.0016) | -1.36 | 0.179 |
| SLF-L | 0.0028 (0.0015) | 0.0025 (0.0018) | 0.48 | 0.6314 |
| SLF-R | 0.0029 (0.0016) | 0.0028 (0.0019) | 0.24 | 0.809 |
| Perioperative Trajectory |  |  |  |  |
| CC Genu | 0.0021 (0.0156) | 0.0011 (0.018) | 0.26 | 0.7961 |
| CC Body | -0.0037 (0.0132) | -0.0008 (0.0203) | -0.71 | 0.4799 |
| CC Splenium | -0.0006 (0.0149) | 0.0001 (0.018) | -0.17 | 0.8672 |
| CST-L | 0.0047 (0.0227) | 0.0031 (0.0188) | 0.31 | 0.7568 |
| CST-R | 0.0038 (0.0233) | -0.0052 (0.0223) | 1.52 | 0.1332 |
| FOF-L | 0.0035 (0.0096) | 0.0042 (0.0197) | -0.19 | 0.8465 |
| FOF-R | 0.0015 (0.0082) | 0.0035 (0.0185) | -0.55 | 0.5846 |
| ILF-L | 0.0051 (0.0106) | 0.0054 (0.018) | -0.1 | 0.9175 |
| ILF-R | 0.0016 (0.0085) | 0.0027 (0.0179) | -0.3 | 0.7638 |
| SLF-L | -0.0017 (0.0179) | 0.0021 (0.0315) | -0.46 | 0.6511 |
| SLF-R | -0.0003 (0.0222) | -0.0004 (0.0239) | 0.01 | 0.9931 |

##### eTable 6c: DTI RD Trajectory differences between the groups with and without WMI

|  | Without WMI  (N=16) | With WMI  (N=26) |  |  |
| --- | --- | --- | --- | --- |
| Radial Diffusivity Trajectories ((10-3mm2/s) /week) | Mean (SD) | Mean (SD) | t-value | p-value |
| Early Infant Trajectory |  |  |  |  |
| CC Genu | -0.0126 (0.0041) | -0.0138 (0.0043) | 1.12 | 0.2677 |
| CC Body | -0.0083 (0.0057) | -0.0111 (0.0064) | 1.82 | 0.0739 |
| CC Splenium | -0.0086 (0.0074) | -0.0109 (0.0062) | 1.3 | 0.1971 |
| CST-L | -0.0095 (0.0054) | -0.011 (0.0058) | 1.01 | 0.315 |
| CST-R | -0.0111 (0.0104) | -0.0118 (0.0101) | 0.25 | 0.8057 |
| FOF-L | -0.0105 (0.0035) | -0.0129 (0.0038) | 2.44 | **0.0181** |
| FOF-R | -0.0128 (0.005) | -0.0137 (0.0033) | 0.78 | 0.4416 |
| ILF-L | -0.0147 (0.0054) | -0.0152 (0.0069) | 0.29 | 0.7762 |
| ILF-R | -0.0148 (0.0049) | -0.0158 (0.0039) | 0.84 | 0.4039 |
| SLF-L | -0.0114 (0.0063) | -0.0098 (0.0053) | -0.9 | 0.3712 |
| SLF-R | -0.0121 (0.0074) | -0.0127 (0.0058) | 0.25 | 0.807 |
| Post-Surgical Trajectory |  |  |  |  |
| CC Genu | -0.0128 (0.0048) | -0.0148 (0.0049) | 1.61 | 0.1118 |
| CC Body | -0.0111 (0.0058) | -0.0135 (0.0066) | 1.52 | 0.1349 |
| CC Splenium | -0.0095 (0.0059) | -0.0111 (0.0058) | 1.08 | 0.2828 |
| CST-L | -0.0084 (0.0068) | -0.0123 (0.0064) | 2.16 | **0.0357** |
| CST-R | -0.0098 (0.0063) | -0.0145 (0.0072) | 2.42 | **0.0192** |
| FOF-L | -0.0114 (0.0054) | -0.0129 (0.005) | 1.1 | 0.2745 |
| FOF-R | -0.0126 (0.0052) | -0.0138 (0.0044) | 0.92 | 0.363 |
| ILF-L | -0.0151 (0.0066) | -0.0157 (0.0067) | 0.35 | 0.7293 |
| ILF-R | -0.0146 (0.0052) | -0.0168 (0.0047) | 1.61 | 0.1136 |
| SLF-L | -0.0156 (0.0064) | -0.0097 (0.0042) | -3.24 | **0.0027** |
| SLF-R | -0.0125 (0.0074) | -0.0113 (0.0088) | -0.45 | 0.6568 |
| Perioperative Trajectory |  |  |  |  |
| CC Genu | -0.0058 (0.0342) | -0.0035 (0.0462) | -0.24 | 0.8073 |
| CC Body | 0.0301 (0.0642) | 0.006 (0.1001) | 1.24 | 0.22 |
| CC Splenium | 0.012 (0.0577) | -0.0103 (0.103) | 1.16 | 0.2515 |
| CST-L | -0.0109 (0.1241) | 0.0083 (0.0646) | -0.78 | 0.4365 |
| CST-R | -0.0104 (0.1658) | 0.0342 (0.1404) | -1.12 | 0.2671 |
| FOF-L | -0.0016 (0.0617) | -0.0154 (0.0557) | 0.95 | 0.344 |
| FOF-R | -0.0115 (0.0421) | -0.0156 (0.0478) | 0.36 | 0.7203 |
| ILF-L | 0.0001 (0.0769) | 0.0131 (0.0748) | -0.71 | 0.4829 |
| ILF-R | -0.0122 (0.0506) | -0.0097 (0.0654) | -0.17 | 0.8621 |
| SLF-L | 0.0394 (0.0829) | -0.0085 (0.0956) | 1.65 | 0.1071 |
| SLF-R | 0.0046 (0.0871) | -0.0067 (0.116) | 0.28 | 0.7842 |

##### eTable 6d. NDT difference between those with WMI and those without WMI

|  | Without WMI | With WMI |  |  |
| --- | --- | --- | --- | --- |
|  | Mean (SD) | Mean (SD) | t-value | p-value |
| 5 Year NDT | (N=16) | (N=26) |  |  |
| WPPSI-III Verbal IQ | 96.438 (19.404) | 94.4 (12.881) | 0.41 | 0.6877 |
| WPPSI-III Full Scale IQ | 98.5 (21.068) | 95.25 (15.369) | 0.56 | 0.5757 |
| Beery-VMI Motor | 86.563 (16.309) | 85.12 (13.293) | 0.31 | 0.7581 |
| 3 Year NDT | (N=13) | (N=32) |  |  |
| Bayley-III Language | 98.615 (10.508) | 93.516 (13.239) | 1.23 | 0.2245 |
| Bayley-III Cognitive | 101 (11.958) | 95.645 (9.376) | 1.59 | 0.1189 |
| Bayley-III Motor | 103.1 (12.692) | 97.484 (12.258) | 1.37 | 0.1789 |
| 1 Year NDT | (N=23) | (N=37) |  |  |
| Bayley-III Language | 90.783 (13.879) | 86.222 (11.492) | 1.37 | 0.176 |
| Bayley-III Cognitive | 103 (16.702) | 100.1 (11.148) | 0.81 | 0.4214 |
| Bayley-III Motor | 91.826 (16.822) | 87.417 (12.159) | 1.17 | 0.2477 |

#### Missing NDT Data Results

The results of the comparison analysis for imaging trajectory differences between participants with and without NDT are presented in eTables 7a, 7b, and 7c (volumes) and eTables 8a, 8b, and 8c (diffusion tractography metrics). Cortex, cerebellum, white matter, right CST FA, left ILF FA and RD, left FOF FA, left SLF RD, and Splenium RD were found to be significantly different between the two groups. However, none of these in their respective imaging epochs were predictive of NDT outcomes at the respective neuropsychological testing time points. The only significant finding of consequence to the main analysis of this study was that deep GM volume trajectory of participants with early childhood NDT testing (both 3-year and 5-year) were higher than participants without NDT testing (Table 7a). This finding was of consequence because the deep GM trajectory was predictive of Language performance at 3 Year in the main multi-variable regression analysis.

##### eTable 7a: Early Infant Volume Trajectory Differences between Groups with and without NDT

|  | 1 Year NDT | | | | 3 Year NDT | | | | 5 Year NDT | | | |
| --- | --- | --- | --- | --- | --- | --- | --- | --- | --- | --- | --- | --- |
|  | Without NDT  (N=25) | With NDT  (N=54) |  |  | Without NDT  (N=38) | With NDT  (N=39) |  |  | Without NDT  (N=37) | With NDT  (N=36) |  |  |
|  | Mean (SD) | Mean (SD) | t-value | p-value | Mean (SD) | Mean (SD) | t-value | p-value | Mean (SD) | Mean (SD) | t-value | p-value |
| Brainstem | 184 (76) | 177 (51) | 0.3745 | 0.7093 | 171 (61) | 183 (52) | 0.8768 | 0.3838 | 172 (61) | 183 (52) | 0.7426 | 0.4604 |
| Cerebellum | 1487 (545) | 1571 (482) | 0.5761 | 0.5665 | 1485 (504) | 1608 (478) | 1.0239 | 0.3096 | 1481 (452) | 1611 (512) | 1.0867 | 0.2811 |
| Cortex | 6801 (4022) | 8702 (2684) | 2.1079 | **0.0388** | 7499 (3359) | 8957 (2663) | 1.993 | 0.0504 | 7699 (3620) | 8817 (2491) | 1.5096 | 0.1359 |
| CSF | 5513 (7306) | 3823 (3257) | 1.3614 | 0.178 | 3840 (5287) | 4150 (3761) | 0.2824 | 0.7785 | 3912 (5338) | 4099 (3714) | 0.1704 | 0.8652 |
| DGM | 846 (614) | 867 (384) | 0.1785 | 0.8589 | 735 (438) | 957 (404) | 2.1578 | **0.0346** | 721 (441) | 967 (396) | 2.4104 | **0.0187** |
| WM | 4812 (3394) | 4761 (1892) | 0.1047 | 0.9169 | 4672 (2604) | 4810 (1935) | 0.2514 | 0.8023 | 4627 (2812) | 4842 (1718) | 0.39 | 0.6978 |
| Whole Brain with CSF | 17527 (3846) | 18038 (3469) | 0.4382 | 0.6627 | 17434 (3108) | 18247 (3755) | 0.9413 | 0.35 | 17325 (3368) | 18323 (3577) | 1.16 | 0.2502 |
| Whole Brain without CSF | 12014 (7099) | 14216 (4015) | 1.5774 | 0.1195 | 13594 (5575) | 14097 (4277) | 0.4211 | 0.675 | 13412 (6004) | 14224 (3832) | 0.6805 | 0.4986 |

##### eTable 7b: Post-Surgical Period Volume Trajectory Differences between Groups with and without NDT

|  | 1 Year NDT | | | | 3 Year NDT | | | | 5 Year NDT | | | |
| --- | --- | --- | --- | --- | --- | --- | --- | --- | --- | --- | --- | --- |
|  | Without NDT  (N=25) | With NDT  (N=50) |  |  | Without NDT  (N=25) | With NDT  (N=37) |  |  | Without NDT  (N=25) | With NDT  (N=36) |  |  |
|  | Mean (SD) | Mean (SD) | t-value | p-value | Mean (SD) | Mean (SD) | t-value | p-value | Mean (SD) | Mean (SD) | t-value | p-value |
| Brainstem | 203 (71) | 200 (65) | 0.1075 | 0.9151 | 175 (70) | 213 (58) | 1.6526 | 0.1085 | 193 (50) | 206 (73) | 0.5702 | 0.5726 |
| Cerebellum | 1387 (701) | 1850 (515) | 1.9124 | 0.0651 | 1467 (605) | 1922 (488) | 2.3334 | **0.0263** | 1570 (587) | 1901 (523) | 1.6913 | 0.1008 |
| Cortex | 6076 (4125) | 9713 (2714) | 2.6469 | **0.0127** | 7905 (4097) | 9521 (2665) | 1.3687 | 0.1809 | 7473 (3984) | 9964 (2258) | 2.2974 | **0.0285** |
| CSF | 4316 (6426) | 4124 (3414) | 0.2258 | 0.8228 | 2607 (4442) | 4651 (3833) | 1.3703 | 0.1805 | 2653 (4106) | 4825 (3954) | 1.5193 | 0.1388 |
| DGM | 1062 (506) | 933 (398) | 0.7169 | 0.4788 | 832 (427) | 1013 (395) | 1.2115 | 0.2349 | 853 (392) | 1017 (416) | 1.1316 | 0.2665 |
| WM | 3802 (3133) | 5881 (2315) | 1.85 | 0.0739 | 3531 (2741) | 6436 (1795) | 3.6649 | **0.0009** | 4219 (2969) | 6280 (1855) | 2.462 | **0.0196** |
| Whole Brain with CSF | 18488 (5384) | 20433 (3164) | 1.1579 | 0.2558 | 17879 (4112) | 21093 (2854) | 2.6275 | **0.0133** | 18204 (3695) | 21203 (3088) | 2.5233 | **0.017** |
| Whole Brain without CSF | 14172 (7409) | 16308 (3901) | 1.0924 | 0.2831 | 15272 (6034) | 16442 (3934) | 0.672 | 0.5066 | 15550 (5530) | 16377 (4147) | 0.4906 | 0.6272 |

##### eTable 7c: Perioperative Period Volume Trajectory Differences between Groups with and without NDT

|  | Year 1 | | | | Year 3 | | | | Year 5 | | | |
| --- | --- | --- | --- | --- | --- | --- | --- | --- | --- | --- | --- | --- |
|  | Without NDT  (N=25) | With NDT  (N=48) |  |  | Without NDT  (N=25) | With NDT  (N=36) |  |  | Without NDT  (N=25) | With NDT  (N=36) |  |  |
|  | Mean (SD) | Mean (SD) | t-value | p-value | Mean (SD) | Mean (SD) | t-value | p-value | Mean (SD) | Mean (SD) | t-value | p-value |
| Brainstem | 166 (541) | 93 (567) | 0.4292 | 0.6704 | 252 (644) | -10 (458) | 1.4679 | 0.1508 | 34 (529) | 160 (578) | 0.7007 | 0.488 |
| Cerebellum | 1235 (2041) | -92 (2392) | 1.6728 | 0.103 | 1198 (2689) | -563 (1862) | 2.3877 | **0.0223** | 865 (2843) | -439 (1725) | 1.7292 | 0.0923 |
| Cortex | 7171 (21066) | 5447 (33022) | 0.0178 | 0.9859 | 9139 (33150) | 6001 (29748) | 0.306 | 0.7614 | 14967 (32793) | 443 (27984) | 1.473 | 0.1495 |
| CSF | 235 (28327) | 5867 (30933) | 0.3043 | 0.7627 | 11450 (34389) | -3443 (27405) | 1.4857 | 0.1461 | 7660 (32702) | -1522 (29547) | 0.9094 | 0.3692 |
| DGM | -1100 (5115) | 608 (3671) | 1.2428 | 0.222 | -529 (4770) | 882 (3623) | 1.0376 | 0.3064 | -80 (4735) | 619 (3624) | 0.5139 | 0.6104 |
| WM | 481 (27483) | -832 (21705) | 0.2314 | 0.8183 | 5975 (25724) | -6032 (19698) | 1.6309 | 0.1116 | 3570 (27921) | -5069 (16936) | 1.1667 | 0.251 |
| Whole Brain with CSF | 12408 (18710) | 7823 (22258) | 0.7389 | 0.4648 | 17362 (21389) | 1152 (20170) | 2.3849 | **0.0225** | 15616 (21790) | 1103 (20225) | 2.1293 | **0.0401** |
| Whole Brain without CSF | 12173 (24389) | 1956 (35529) | 0.791 | 0.4341 | 5912 (33240) | 4595 (32811) | 0.1215 | 0.904 | 7955 (37343) | 2624 (28305) | 0.499 | 0.6208 |

##### eTable 8a: Early Infant Period DTI (FA & RD) Trajectory Differences between Groups with and without NDT

|  | 1 Year NDT | | | | 3 Year NDT | | | | 5 Year NDT | | | |
| --- | --- | --- | --- | --- | --- | --- | --- | --- | --- | --- | --- | --- |
|  | Without NDT  (N=25) | With NDT  (N=54) |  |  | Without NDT  (N=38) | With NDT  (N=39) |  |  | Without NDT  (N=37) | With NDT  (N=36) |  |  |
|  | Mean (SD) | Mean (SD) | t-value | p-value | Mean (SD) | Mean (SD) | t-value | p-value | Mean (SD) | Mean (SD) | t-value | p-value |
| FA |  |  |  |  |  |  |  |  |  |  |  |  |
| CCBody | 0.0022 (0.0009) | 0.0024 (0.0011) | 0.5592 | 0.5782 | 0.0022 (0.0009) | 0.0024 (0.0012) | 0.893 | 0.3755 | 0.0021 (0.0009) | 0.0025 (0.0011) | 1.559 | 0.1243 |
| CST-L | 0.0044 (0.002) | 0.0047 (0.0015) | 0.4496 | 0.6548 | 0.0045 (0.0016) | 0.0047 (0.0015) | 0.5091 | 0.6128 | 0.0046 (0.0016) | 0.0047 (0.0016) | 0.0721 | 0.9428 |
| CST-R | 0.0052 (0.0024) | 0.0047 (0.0018) | 0.7941 | 0.4307 | 0.0051 (0.0019) | 0.0046 (0.0019) | 0.8338 | 0.4081 | 0.0051 (0.0018) | 0.0046 (0.0019) | 0.9716 | 0.3356 |
| FOF-L | 0.0039 (0.0013) | 0.003 (0.0011) | 2.3172 | **0.0242** | 0.0037 (0.0012) | 0.0029 (0.0011) | 2.6478 | **0.0105** | 0.0036 (0.0012) | 0.003 (0.0011) | 1.9569 | 0.0554 |
| FOF-R | 0.0034 (0.0017) | 0.0031 (0.0012) | 0.645 | 0.522 | 0.0035 (0.0013) | 0.0029 (0.0013) | 1.4469 | 0.1544 | 0.0033 (0.0013) | 0.0031 (0.0013) | 0.7313 | 0.4682 |
| Genu | 0.0029 (0.0019) | 0.0034 (0.001) | 1.2757 | 0.2072 | 0.0031 (0.0015) | 0.0034 (0.0011) | 0.8246 | 0.413 | 0.0033 (0.0015) | 0.0033 (0.0011) | 0.0779 | 0.9382 |
| ILF-L | 0.0043 (0.0009) | 0.0038 (0.0013) | 1.4117 | 0.1637 | 0.0041 (0.001) | 0.0037 (0.0013) | 1.3262 | 0.1902 | 0.0041 (0.001) | 0.0038 (0.0014) | 0.9129 | 0.3653 |
| ILF-R | 0.0032 (0.0016) | 0.0036 (0.0013) | 0.843 | 0.4029 | 0.0035 (0.0016) | 0.0035 (0.0012) | 0.1761 | 0.8609 | 0.0034 (0.0014) | 0.0036 (0.0014) | 0.5553 | 0.581 |
| SLF-L | 0.0021 (0.0019) | 0.0023 (0.0012) | 0.5548 | 0.582 | 0.0022 (0.0016) | 0.0024 (0.001) | 0.5262 | 0.6015 | 0.0023 (0.0016) | 0.0022 (0.0011) | 0.2962 | 0.7686 |
| SLF-R | 0.0026 (0.0016) | 0.0029 (0.0016) | 0.5804 | 0.566 | 0.0027 (0.0013) | 0.003 (0.0019) | 0.6549 | 0.5175 | 0.0025 (0.0014) | 0.0032 (0.0018) | 1.192 | 0.2426 |
| Splenium | 0.0025 (0.0026) | 0.0031 (0.0014) | 1.1677 | 0.2476 | 0.0032 (0.0018) | 0.0028 (0.0017) | 0.8432 | 0.4025 | 0.0028 (0.0019) | 0.0032 (0.0016) | 0.9386 | 0.3518 |
| RD |  |  |  |  |  |  |  |  |  |  |  |  |
| CCBody | -0.0076 (0.0077) | -0.0102 (0.0056) | 1.3537 | 0.181 | -0.0096 (0.0077) | -0.0097 (0.0046) | 0.0945 | 0.925 | -0.0093 (0.0062) | -0.0099 (0.0062) | 0.4304 | 0.6685 |
| CST-L | -0.0109 (0.0071) | -0.0101 (0.0053) | 0.4126 | 0.6816 | -0.0105 (0.005) | -0.01 (0.006) | 0.3008 | 0.7648 | -0.01 (0.0054) | -0.0104 (0.0058) | 0.2758 | 0.7838 |
| CST-R | -0.0137 (0.0101) | -0.0109 (0.0102) | 0.7806 | 0.4385 | -0.0119 (0.0077) | -0.0111 (0.0116) | 0.2922 | 0.7713 | -0.0119 (0.008) | -0.0111 (0.0115) | 0.2839 | 0.7776 |
| FOF-L | -0.0108 (0.0045) | -0.0119 (0.0036) | 0.8793 | 0.3831 | -0.0113 (0.0042) | -0.0119 (0.0036) | 0.5366 | 0.5937 | -0.0115 (0.004) | -0.0118 (0.0037) | 0.2443 | 0.8079 |
| FOF-R | -0.0134 (0.0067) | -0.0132 (0.0034) | 0.1806 | 0.8574 | -0.0139 (0.0053) | -0.0127 (0.0032) | 1.0036 | 0.3206 | -0.0143 (0.0049) | -0.0124 (0.0034) | 1.5989 | 0.1164 |
| Genu | -0.0128 (0.0038) | -0.0133 (0.0044) | 0.3626 | 0.7182 | -0.013 (0.0046) | -0.0133 (0.004) | 0.3061 | 0.7606 | -0.0135 (0.0032) | -0.0129 (0.0049) | 0.5424 | 0.5896 |
| ILF-L | -0.0159 (0.0068) | -0.0147 (0.0059) | 0.6143 | 0.5416 | -0.0152 (0.006) | -0.0147 (0.0063) | 0.2807 | 0.78 | -0.0157 (0.0056) | -0.0143 (0.0065) | 0.8465 | 0.401 |
| ILF-R | -0.0159 (0.0063) | -0.0152 (0.0039) | 0.5081 | 0.6134 | -0.0156 (0.0051) | -0.0152 (0.004) | 0.34 | 0.7352 | -0.0154 (0.0048) | -0.0153 (0.0043) | 0.1029 | 0.9184 |
| SLF-L | -0.0104 (0.0089) | -0.0106 (0.0047) | 0.1314 | 0.896 | -0.0115 (0.0066) | -0.0095 (0.0047) | 1.1284 | 0.2656 | -0.0116 (0.0066) | -0.0097 (0.0048) | 1.0971 | 0.2789 |
| SLF-R | -0.0127 (0.0102) | -0.0122 (0.0053) | 0.1778 | 0.86 | -0.0121 (0.008) | -0.0126 (0.0055) | 0.2343 | 0.8163 | -0.0115 (0.0082) | -0.0133 (0.0049) | 0.7476 | 0.4605 |
| Splenium | -0.0058 (0.0102) | -0.0108 (0.0053) | 2.4251 | **0.0184** | -0.0096 (0.0088) | -0.0098 (0.0051) | 0.0869 | 0.9311 | -0.0084 (0.0078) | -0.0106 (0.0061) | 1.2421 | 0.2191 |

##### eTable 8b: Post-Surgical Period DTI Trajectory Differences between Groups with and without NDT

|  | 1 Year NDT | | | | 3 Year NDT | | | | 5 Year NDT | | | |
| --- | --- | --- | --- | --- | --- | --- | --- | --- | --- | --- | --- | --- |
|  | Without NDT  (N=25) | With NDT  (N=50) |  |  | Without NDT  (N=25) | With NDT  (N=37) |  |  | Without NDT  (N=25) | With NDT  (N=36) |  |  |
|  | Mean (SD) | Mean (SD) | t-value |  | Mean (SD) | Mean (SD) | t-value |  | Mean (SD) | Mean (SD) | t-value |  |
| FA |  |  |  |  |  |  |  |  |  |  |  |  |
| CCBody | 0.0029 (0.0014) | 0.0028 (0.0011) | 0.1099 | 0.9129 | 0.0026 (0.0012) | 0.003 (0.0011) | 1.1665 | 0.248 | 0.0027 (0.0012) | 0.003 (0.0011) | 0.9454 | 0.3482 |
| CST-L | 0.0047 (0.0018) | 0.0046 (0.0017) | 0.2734 | 0.7856 | 0.0047 (0.0017) | 0.0045 (0.0017) | 0.4918 | 0.6248 | 0.0045 (0.002) | 0.0046 (0.0015) | 0.2122 | 0.8328 |
| CST-R | 0.0062 (0.0024) | 0.0051 (0.0021) | 1.2282 | 0.2252 | 0.0053 (0.0025) | 0.0053 (0.0021) | 0.0747 | 0.9408 | 0.0056 (0.0025) | 0.0051 (0.002) | 0.7569 | 0.4527 |
| FOF-L | 0.0035 (0.0014) | 0.0031 (0.0013) | 0.8607 | 0.3929 | 0.0035 (0.0015) | 0.003 (0.0012) | 1.4283 | 0.1586 | 0.0034 (0.0014) | 0.0031 (0.0013) | 0.8301 | 0.4099 |
| FOF-R | 0.0038 (0.0015) | 0.0031 (0.0011) | 1.5155 | 0.1356 | 0.0036 (0.0014) | 0.003 (0.001) | 1.7059 | 0.0939 | 0.0033 (0.0013) | 0.0032 (0.001) | 0.1773 | 0.8599 |
| Genu | 0.003 (0.0016) | 0.0036 (0.0013) | 1.321 | 0.1914 | 0.0031 (0.0015) | 0.0037 (0.0012) | 1.574 | 0.1207 | 0.0035 (0.0015) | 0.0034 (0.0012) | 0.0746 | 0.9408 |
| ILF-L | 0.0045 (0.0014) | 0.0035 (0.0014) | 2.2424 | **0.0288** | 0.0041 (0.0016) | 0.0034 (0.0013) | 1.8393 | 0.071 | 0.0041 (0.0015) | 0.0033 (0.0014) | 2.0276 | **0.0472** |
| ILF-R | 0.0035 (0.0012) | 0.0036 (0.0014) | 0.1353 | 0.8929 | 0.0035 (0.0015) | 0.0037 (0.0014) | 0.3473 | 0.7297 | 0.0035 (0.0013) | 0.0037 (0.0015) | 0.423 | 0.6739 |
| SLF-L | 0.0029 (0.002) | 0.0025 (0.0015) | 0.6285 | 0.534 | 0.0024 (0.0018) | 0.0028 (0.0014) | 0.7169 | 0.4785 | 0.0027 (0.0019) | 0.0026 (0.0013) | 0.1095 | 0.9135 |
| SLF-R | 0.0018 (0.0012) | 0.0032 (0.0017) | 1.9341 | 0.0623 | 0.0026 (0.0017) | 0.0031 (0.0017) | 0.7175 | 0.4784 | 0.0028 (0.0017) | 0.003 (0.0017) | 0.4129 | 0.6825 |
| Splenium | 0.0029 (0.0024) | 0.0033 (0.0013) | 0.9023 | 0.3705 | 0.0034 (0.0016) | 0.0031 (0.0015) | 0.7851 | 0.4354 | 0.0031 (0.0017) | 0.0033 (0.0015) | 0.4299 | 0.6687 |
| RD |  |  |  |  |  |  |  |  |  |  |  |  |
| CCBody | -0.0128 (0.0086) | -0.012 (0.0055) | 0.4368 | 0.6638 | -0.0122 (0.0072) | -0.0121 (0.0054) | 0.0188 | 0.985 | -0.0132 (0.0067) | -0.0113 (0.0056) | 1.2119 | 0.2302 |
| CST-L | -0.0107 (0.0075) | -0.01 (0.0067) | 0.2943 | 0.7697 | -0.0104 (0.006) | -0.01 (0.0073) | 0.2399 | 0.8113 | -0.0101 (0.0065) | -0.0102 (0.0071) | 0.0575 | 0.9544 |
| CST-R | -0.0145 (0.0085) | -0.0114 (0.0067) | 1.1359 | 0.2615 | -0.0121 (0.0068) | -0.0118 (0.0072) | 0.1116 | 0.9116 | -0.0123 (0.0064) | -0.0117 (0.0074) | 0.3306 | 0.7423 |
| FOF-L | -0.0132 (0.008) | -0.0117 (0.0043) | 0.921 | 0.3609 | -0.0126 (0.0064) | -0.0116 (0.0043) | 0.6622 | 0.5105 | -0.0126 (0.0061) | -0.0115 (0.0044) | 0.818 | 0.4167 |
| FOF-R | -0.0131 (0.008) | -0.013 (0.0041) | 0.0647 | 0.9487 | -0.0131 (0.0059) | -0.013 (0.0042) | 0.1017 | 0.9194 | -0.0132 (0.0056) | -0.0129 (0.0043) | 0.1761 | 0.8609 |
| Genu | -0.0143 (0.0054) | -0.0135 (0.0048) | 0.4851 | 0.6294 | -0.0136 (0.0049) | -0.0137 (0.005) | 0.1122 | 0.911 | -0.0147 (0.0044) | -0.0128 (0.0052) | 1.4987 | 0.1391 |
| ILF-L | -0.0182 (0.0067) | -0.0146 (0.0064) | 1.7331 | 0.0884 | -0.0167 (0.0066) | -0.0144 (0.0064) | 1.3847 | 0.1714 | -0.0174 (0.0065) | -0.0136 (0.0062) | 2.3629 | **0.0215** |
| ILF-R | -0.0165 (0.0074) | -0.0152 (0.0045) | 0.7518 | 0.4554 | -0.0146 (0.006) | -0.016 (0.0044) | 1.0244 | 0.3102 | -0.0155 (0.0058) | -0.0154 (0.0046) | 0.0477 | 0.9622 |
| SLF-L | -0.0137 (0.0076) | -0.0122 (0.0056) | 0.6624 | 0.5123 | -0.0129 (0.0071) | -0.0123 (0.0052) | 0.3088 | 0.7594 | -0.0147 (0.0071) | -0.0106 (0.0042) | 2.1053 | **0.043** |
| SLF-R | -0.0114 (0.0108) | -0.0121 (0.0073) | 0.213 | 0.8327 | -0.0112 (0.0108) | -0.0126 (0.0049) | 0.5109 | 0.613 | -0.013 (0.0075) | -0.0107 (0.0087) | 0.8159 | 0.4208 |
| Splenium | -0.0074 (0.0088) | -0.0109 (0.0046) | 2.0055 | **0.0494** | -0.0104 (0.0067) | -0.0101 (0.0052) | 0.187 | 0.8523 | -0.0108 (0.0064) | -0.0097 (0.0053) | 0.7013 | 0.4858 |

##### eTable 8c: Perioperative Period DTI Trajectory Differences between Groups with and without NDT

|  | 1 Year NDT | | | | 3 Year NDT | | | | 5 Year NDT | | | |
| --- | --- | --- | --- | --- | --- | --- | --- | --- | --- | --- | --- | --- |
|  | Without NDT  (N=25) | With NDT  (N=48) |  |  | Without NDT  (N=25) | With NDT  (N=36) |  |  | Without NDT  (N=25) | With NDT  (N=36) |  |  |
|  | Mean (SD) | Mean (SD) | t-value | p-value | Mean (SD) | Mean (SD) | t-value | p-value | Mean (SD) | Mean (SD) | t-value | p-value |
| FA |  |  |  |  |  |  |  |  |  |  |  |  |
| CCBody | -0.0003 (0.0173) | -0.0033 (0.0169) | 0.7008 | 0.4857 | -0.0016 (0.019) | -0.003 (0.0147) | 0.3404 | 0.7346 | -0.0019 (0.018) | -0.0027 (0.0162) | 0.1955 | 0.8456 |
| CST-L | 0.0049 (0.0153) | 0.0035 (0.0231) | 0.2553 | 0.7993 | 0.0049 (0.0218) | 0.003 (0.02) | 0.3763 | 0.7079 | 0.0051 (0.0206) | 0.0027 (0.0212) | 0.4815 | 0.6317 |
| CST-R | 0.0009 (0.0196) | -0.0012 (0.0246) | 0.3094 | 0.7581 | 0.0034 (0.0222) | -0.0036 (0.0236) | 1.1608 | 0.2505 | 0.0079 (0.0207) | -0.0066 (0.023) | 2.5192 | **0.0145** |
| FOF-L | 0.0051 (0.0083) | 0.0032 (0.0176) | 0.4743 | 0.6368 | 0.0041 (0.0195) | 0.0036 (0.0095) | 0.1549 | 0.8774 | 0.0061 (0.0186) | 0.0015 (0.0102) | 1.2298 | 0.2232 |
| FOF-R | 0.0018 (0.0085) | 0.0028 (0.0161) | 0.2822 | 0.7788 | 0.0031 (0.0181) | 0.0018 (0.0081) | 0.3849 | 0.7017 | 0.004 (0.0175) | 0.0007 (0.008) | 0.9137 | 0.3645 |
| Genu | 0.0033 (0.0152) | 0.0007 (0.0175) | 0.6196 | 0.5375 | 0.0051 (0.0185) | -0.0019 (0.0139) | 1.819 | 0.0731 | 0.0021 (0.0169) | 0.0011 (0.0166) | 0.2367 | 0.8135 |
| ILF-L | 0.0036 (0.0101) | 0.0061 (0.0165) | 0.6729 | 0.5034 | 0.0051 (0.017) | 0.0054 (0.0119) | 0.0913 | 0.9276 | 0.0045 (0.0171) | 0.006 (0.0118) | 0.4384 | 0.6625 |
| ILF-R | -0.0012 (0.0106) | 0.0037 (0.015) | 1.3428 | 0.1842 | 0.0031 (0.0177) | 0.0011 (0.0085) | 0.5882 | 0.5585 | 0.0023 (0.0169) | 0.0019 (0.01) | 0.0982 | 0.9221 |
| SLF-L | -0.0031 (0.0162) | 0.0018 (0.0287) | 0.5698 | 0.5723 | 0.0052 (0.0268) | -0.0063 (0.0216) | 1.4287 | 0.1617 | 0.0037 (0.0278) | -0.0039 (0.0215) | 0.9337 | 0.3567 |
| SLF-R | -0.0049 (0.0204) | 0.0011 (0.0233) | 0.5701 | 0.5742 | -0.0065 (0.0244) | 0.0064 (0.0188) | 1.4697 | 0.1552 | -0.006 (0.0218) | 0.0118 (0.0197) | 1.9606 | 0.0621 |
| Splenium | -0.0014 (0.0142) | 0.0003 (0.0175) | 0.4354 | 0.6646 | -0.0012 (0.0132) | 0.0007 (0.0193) | 0.4854 | 0.6289 | -0.0008 (0.0154) | 0.0002 (0.0175) | 0.2489 | 0.8041 |
| RD |  |  |  |  |  |  |  |  |  |  |  |  |
| CCBody | 0.0186 (0.0815) | 0.0183 (0.086) | 0.0137 | 0.9891 | 0.016 (0.1117) | 0.0209 (0.0386) | 0.2534 | 0.8007 | 0.0271 (0.0732) | 0.0097 (0.0936) | 0.8885 | 0.3772 |
| CST-L | -0.0101 (0.0768) | 0.0021 (0.1104) | 0.4648 | 0.6436 | -0.0145 (0.0533) | 0.01 (0.1299) | 1.0057 | 0.3182 | -0.0004 (0.0634) | -0.0034 (0.1296) | 0.1228 | 0.9026 |
| CST-R | 0.0147 (0.1977) | 0.0096 (0.1345) | 0.1163 | 0.9078 | -0.0121 (0.1567) | 0.0289 (0.1525) | 1.0191 | 0.3124 | -0.0096 (0.1491) | 0.026 (0.1585) | 0.8808 | 0.382 |
| FOF-L | 0.0074 (0.0735) | -0.0159 (0.0494) | 1.5364 | 0.1293 | 0.0006 (0.0698) | -0.0168 (0.0453) | 1.2147 | 0.2289 | -0.0021 (0.0693) | -0.0145 (0.046) | 0.8658 | 0.3898 |
| FOF-R | -0.0123 (0.0336) | -0.0139 (0.0496) | 0.1397 | 0.8894 | -0.0178 (0.048) | -0.009 (0.041) | 0.775 | 0.4414 | -0.0175 (0.0467) | -0.0086 (0.0422) | 0.7833 | 0.4365 |
| Genu | -0.0089 (0.0404) | -0.0025 (0.0402) | 0.639 | 0.5249 | -0.0093 (0.0462) | 0.0001 (0.0327) | 1.0038 | 0.3189 | -0.005 (0.0457) | -0.0043 (0.034) | 0.0741 | 0.9412 |
| ILF-L | 0.0141 (0.0833) | 0.0025 (0.0721) | 0.5995 | 0.5509 | 0.0143 (0.0912) | -0.0015 (0.0562) | 0.856 | 0.3951 | 0.0026 (0.0821) | 0.0102 (0.0695) | 0.4129 | 0.681 |
| ILF-R | -0.0063 (0.0552) | -0.0133 (0.0595) | 0.4566 | 0.6495 | -0.0199 (0.0729) | -0.0021 (0.0361) | 1.2333 | 0.2221 | -0.0156 (0.0622) | -0.0064 (0.0536) | 0.63 | 0.531 |
| SLF-L | 0.0166 (0.1098) | 0.0168 (0.0825) | 0.0081 | 0.9936 | -0.008 (0.0952) | 0.0473 (0.0783) | 1.9229 | 0.0624 | -0.0041 (0.0951) | 0.0399 (0.0832) | 1.5127 | 0.1391 |
| SLF-R | -0.0417 (0.102) | 0.0132 (0.095) | 1.2143 | 0.2369 | 0.0009 (0.1209) | -0.0009 (0.0693) | 0.045 | 0.9645 | 0.0132 (0.1087) | -0.0279 (0.0654) | 0.9836 | 0.3356 |
| Splenium | -0.0005 (0.0564) | 0.002 (0.0944) | 0.1179 | 0.9064 | -0.0043 (0.1067) | 0.0069 (0.0478) | 0.5738 | 0.5679 | 0.0125 (0.0619) | -0.0102 (0.0995) | 1.1815 | 0.2413 |

##### eTable 9: Sociodemographic Factor Differences between Groups with and without NDT

|  |  | With NDT | Without NDT |
| --- | --- | --- | --- |
|  | p-value | mean (SD) | mean (SD) |
| 5-Year |  | N=36 | N=37 |
| SES | 0.3654 | 41.09 (13.50) | 37.00 (16.65) |
| Maternal IQ | 0.5233 | 104.08 (17.92) | 101.05 (15.10) |
| Latino | 0.4718 | 8 | 10 |
| White | 0.2567 | 26 | 22 |
| Black | 0.2544 | 2 | 5 |
| Asian – only 2 were recruited but did not attend ND testing | | | |
| 3-Year |  | N=39 | N=38 |
| SES | 0.0941 | 41.89 (13.78) | 34.08 (15.35) |
| Maternal IQ | 0.2198 | 104.90 (16.87) | 98.94 (16.75) |
| Latino | 0.3974 | 8 | 11 |
| White | 0.1300 | 29 | 22 |
| Black | 0.2258 | 2 | 5 |
| Asian – only 2 were recruited but did not attend ND testing | | | |
| 1-Year |  | N=54 | N=25 |
| SES | 0.5047 | 39.51 (14.48) | 45.33 (16.04) |
| Maternal IQ | 0.3315 | 102.5 (17.01) | 112.33 (14.36) |
| Latino | 0.8808 | 15 | 6 |
| White | 0.2265 | 32 | 17 |
| Black | 0.9383 | 5 | 2 |
| Asian – only 2 were recruited but did not attend ND testing | | | |

No differences in SES, Maternal IQ, or race/ethnicity in people who returned for testing and who did not return.

##### eTable 10: One-year and Three-year Neurodevelopmental differences between Participants that Attended Five-year Assessment and Those Who Did Not

|  |  | Had 5-yr | No 5-yr |
| --- | --- | --- | --- |
|  | p-value | mean (sd) | mean (sd) |
| 3-Year |  | N=32 | N=5 |
| Bayley-III Language | 0.8242 | 96.19 (13.66) | 97.6 (7.77) |
| Bayley-III Cognitive | 0.3654 | 99.22 (10.56) | 94.6 (9.79) |
| Bayley-III Motor | 0.5634 | 100.66 (12.91) | 97.2 (6.02) |
| 1-Year |  | N=36 | N=18 |
| Bayley-III Language | 0.6483 | 89.73 (11.97) | 91.7 (12.31) |
| Bayley-III Cognitive | 0.0568 | 104.59 (12.93) | 96.36 (9.51) |
| Bayley-III Motor | 0.1184 | 92.7 (12.75) | 85.6 (11.56) |

No differences in NDT at 1-year and 3-year between participants who completed 5-year assessment and participants who did not attend 5-year assessment.
